# Population Genomics of *Plasmodium malariae* from Four African Countries

**DOI:** 10.1101/2024.09.07.24313132

**Authors:** Zachary R. Popkin-Hall, Kelly Carey-Ewend, Farhang Aghakhanian, Eniyou C. Oriero, Misago D. Seth, Melchior M. Kashamuka, Billy Ngasala, Innocent M. Ali, Eric Sompwe Mukomena, Celine I. Mandara, Oksana Kharabora, Rachel Sendor, Alfred Simkin, Alfred Amambua-Ngwa, Antoinette Tshefu, Abebe A. Fola, Deus S. Ishengoma, Jeffrey A. Bailey, Jonathan B. Parr, Jessica T. Lin, Jonathan J. Juliano

## Abstract

*Plasmodium malariae* is geographically widespread but neglected and may become more prevalent as *P. falciparum* declines. We completed the largest genomic study of African *P. malariae* to-date by performing hybrid capture and sequencing of 77 isolates from Cameroon (n=7), the Democratic Republic of the Congo (n=16), Nigeria (n=4), and Tanzania (n=50) collected between 2015 and 2021. There is no evidence of geographic population structure. Nucleotide diversity was significantly lower than in co-localized *P. falciparum* isolates, while linkage disequilibrium was significantly higher. Genome-wide selection scans identified no erythrocyte invasion ligands or antimalarial resistance orthologs as top hits; however, targeted analyses of these loci revealed evidence of selective sweeps around four erythrocyte invasion ligands and six antimalarial resistance orthologs. Demographic inference modeling suggests that African *P. malariae* is recovering from a bottleneck. Altogether, these results suggest that *P. malariae* is genomically atypical among human *Plasmodium* spp. and panmictic in Africa.

---

*Plasmodium malariae* is a neglected malaria parasite species with a broad but irregular global distribution^1^ and the ability to cause persistent infections^2^. Parasite densities are typically lower than *Plasmodium falciparum*^3^ and most infections involve multiple species^3^. While *P. malariae* causes less severe clinical disease than *P. falciparum*, it can still be deadly owing to severe complications such as glomerulonephritis and/or anemia^4–6^. Prevalence estimates in sub-Saharan Africa vary, but may rival *P. falciparum* in some settings^3,7–11^, with a particularly high contribution to malaria morbidity during the dry season in areas with seasonal transmission^12^. Evidence from Tanzania suggests that as control measures reduce *P. falciparum* cases, *P. malariae* may become more prevalent^13^, as has occurred with *P. knowlesi* in Malaysia^14^ and *P. vivax* in the Solomon Islands^15^ and elsewhere^16^.

*P. malariae* is most closely related to non-human primate malarias found throughout African apes, as well as *Plasmodium brasilianum*, which may represent a recent anthroponosis^17^. These species form a distinct clade from other human *Plasmodium* spp.^17^. While the *P. malariae* genome is incomplete, it shares many 1-1 orthologous genes with *P. falciparum*, including genes that are putatively implicated in antimalarial resistance and erythrocyte invasion based on their *P. falciparum* orthologs^18^. Though genetic studies of *P. malariae* are extremely limited and only one incomplete reference genome is available, previous analyses suggest that it experienced a bottleneck after spilling over from non-human apes to humans^17^.

To date, genomic analysis of *P. malariae* has been limited. A recent microsatellite study of 75 *P. malariae* isolates from seven African countries identified no geographic structure, high diversity among the microsatellite markers, and strong linkage disequilibrium (LD)^19^. Microsatellite markers also showed higher diversity in African *P. malariae* than South American and Asian populations^19^. A single previous whole-genome sequencing study of *P. malariae* leveraged selective whole-genome amplification of 18 isolates from sub-Saharan Africa and Thailand. This study identified that the Thai isolates clustered independently from the African isolates in a maximum-likelihood phylogeny^20^. The study also identified mutations in the putative antimalarial resistance genes dihydrofolate reductase (*pmdhfr*), dihydropteroate synthase (*pmdhps*), and multridrug resistance protein 1 (*pmmdr1*)^20^.

To improve understanding of the population genetics and demographic history of *P. malariae*, we conducted the largest genomic analysis of *P. malariae* in Africa to-date, incorporating 77 whole genomes generated using a custom hybrid capture protocol. These 77 isolates span four high-transmission African countries: Cameroon, the Democratic Republic of the Congo, Nigeria, and Tanzania. These genomes were used to characterize the *P. malariae* population in Africa, and compared to *P. falciparum* from similar geographic regions.

## Results

### High sequencing coverage of African *P. malariae* isolates

Of 81 genomic DNA samples that underwent hybrid capture enrichment and sequencing, 77 yielded usable sequences and were used for downstream genomic analysis. These 77 samples include 7 samples from Cameroon, 16 from the Democratic Republic of the Congo, 4 from Nigeria, and 50 from Tanzania. Tanzanian samples spanned 14 regions across Tanzania.Enrichment was overall highly successful, but a lower proportion of *P. malariae* reads were extracted from lower density samples (**Supplemental Figure 1**). In the 77 samples that were successfully enriched and sequenced, sequencing coverage was high, with the mean coverage across all chromosomes surpassing 50-fold (**Supplemental Figure 2**). Forty samples (51.9%) had >50X coverage across all chromosomes, 58 samples (75.3%) had >20X coverage across all chromosomes, and 67 samples (87.0%) had >10X coverage across all chromosomes. A total of 178,179 high-quality SNPs were identified following quality and missingness filtering and repeat masking (see **Methods**).

### Low complexity of infection

Complexity of infection (COI) was estimated using coiaf^21^ and was compared to that of 662 geographically matched *P. falciparum* isolates from the publicly available MalariaGEN *Pf7* dataset ^22^ **(Figure 1)**. The majority (92.2%, n = 71) of *P. malariae* isolates were monoclonal (COI= 1). Among the remaining six polyclonal samples, all but one contained two clones, while the remaining sample was estimated to contain three clones. By contrast, polyclonal infections were significantly (ANOVA F = 12.5, p < 0.001, df = 1) more common in *P. falciparum*. Of 662 geographically matched *P. falciparum* isolates, only 384 (58.0%) were monoclonal, while the remaining 278 (42.0%) were polyclonal. Infection by two clones was the most common form of *P. falciparum* polyclonal infection (71.6%, n = 199), similar to *P. malariae*. While country of origin was significantly associated with COI (F = 2.90, p = 0.034, df = 3), the interaction between species and country was not (F = 1.08, p = 0.340, df = 2). A Tukey post-hoc test identified no significant differences in COI by country within species.

**Figure 1.**
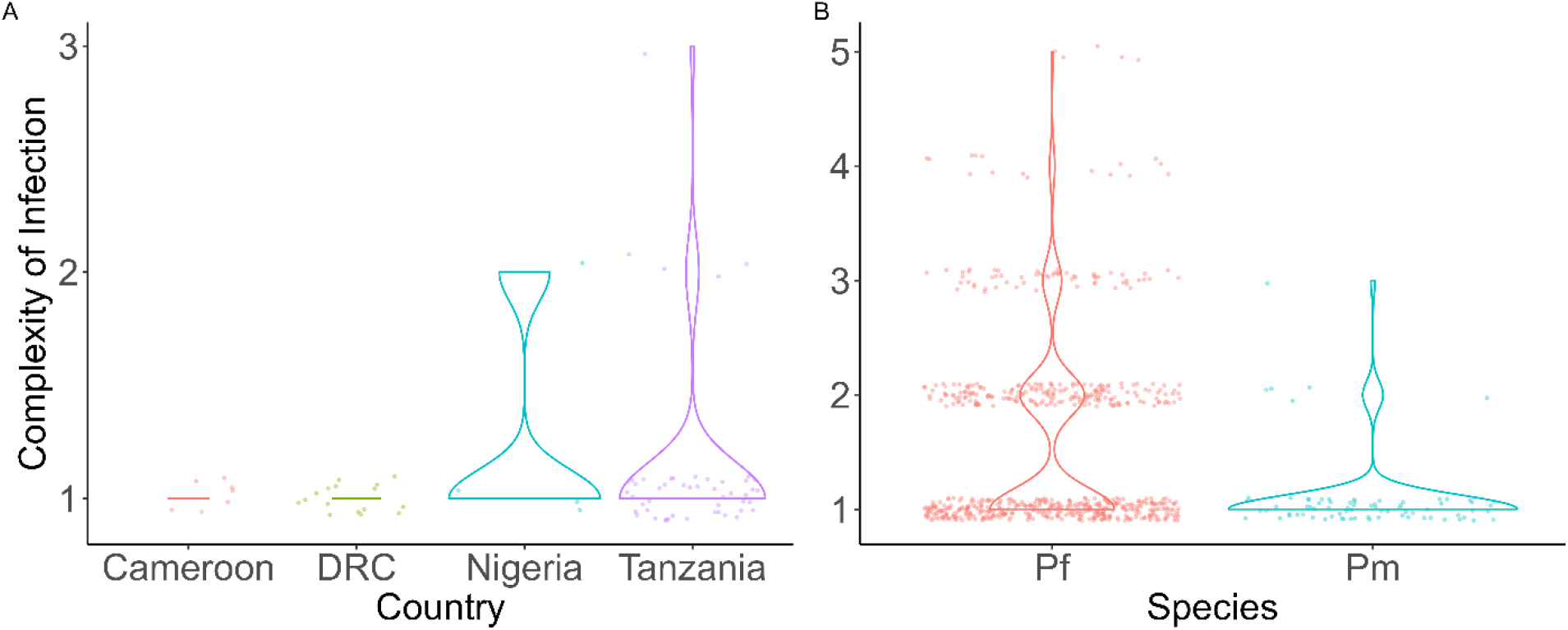
Complexity of infection in A) *P. malariae* by country and B) geographically matched *P. falciparum* and *P. malariae* isolates overall. COI values are significantly lower in *P. malariae* than *P. falciparum* (ANOVA F = 12.5, p < 0.001, df = 1), but there is no significant variation by country within species.

### Low nucleotide diversity

Among 3,763 candidate one-to-one orthologs between *P. malariae* and *P. falciparum*, 1,377 ortholog pairs were retained for nucleotide diversity (π) analysis following masking for coverage and repetitive regions (see **Methods**). The average π in *P. malariae* was 1.72 x 10^-4^, significantly lower (t = -113, p < 0.001, df = 5340) than the average π in *P. falciparum*, which was 6.11 x 10^-3^ (**Figure 2A**). SNP density was also lower in *P. malariae* than *P. falciparum*(**Supplemental Figure 3**).

**Figure 2.**
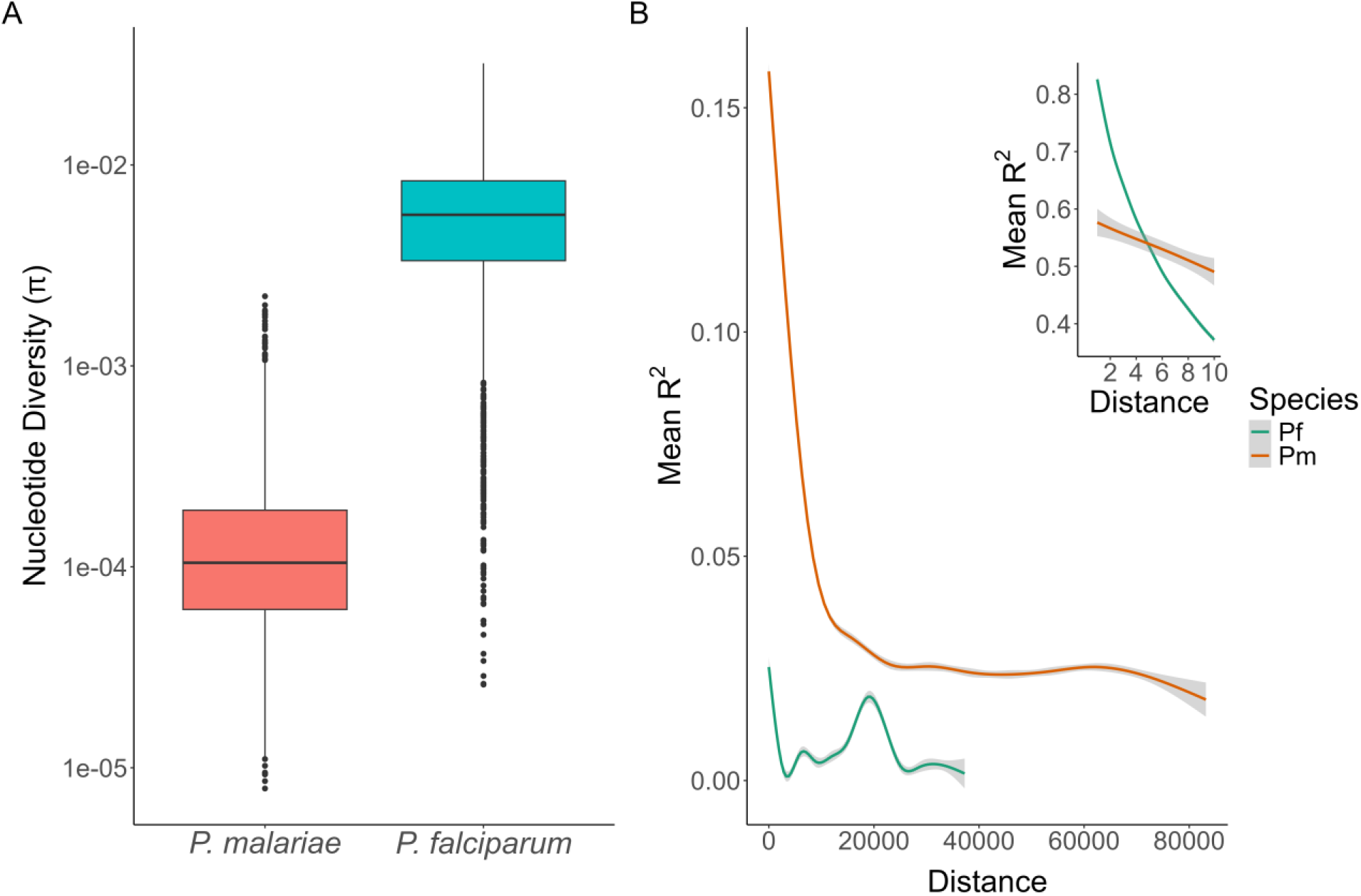
A) Nucleotide diversity (π) of orthologous genes among *P. malariae* and *P. falciparum* isolates. A log-transformed boxplot of π for each gene is shown for each of the 1,377 orthologs retained after masking. 68 orthologs where missing data precluded π calculation for *P. malariae* are not shown. Boxes represent the 25^th^, 50^th^, and 75^th^ percentiles, with outliers represented by dots. The difference in π between the species is highly significant (t= -113, p < 0.001, df = 5340). **B) Linkage disequilibrium (LD) decay in *P. falciparum and P. malariae***. R^2^ values were calculated in PLINK for each distance. LD is higher for *P. malariae* compared to *P. falciparum*, with rapid decay of LD in *P. falciparum* over short distances. The LD decay difference is highly significant (t = -108, p < 0.001, df = 73,920). The insert shows LD from 2 to 10bp, demonstrating that linkage in *P. falciparum* is higher than in *P. malariae* only at very short distances (<8 bp).

### High linkage disequilibrium (LD)

LD was calculated in PLINK^23^ for both *P. malariae* and the geographically matched *P. falciparum* isolates. Mean R^2^ values supporting linkage were plotted by distance between base pairs for both species (**Figure 2B**). While the mean R^2^ is higher in *P. falciparum* for distances < 5 bp (see **Figure 2B inset**), it is consistently higher in *P. malariae* at distances ≥ 5 bp. This difference in LD between the two species is highly significant (t = -108, p < 0.001, df = 73,920).

### No evidence of population structure or geographic differentiation

Principal component analysis (PCA) of the 71 monoclonal *P. malariae* isolates shows no clear structure and no clear relationship between geography and genetic relatedness among isolates, as well as low explanatory value, likely pointing to the lack of genetic variation (**Figure 3**). Discriminant PCA (DAPC) identified seven population clusters, three clusters with multiple samples and four singletons, as the most likely possibility according to the Bayes Information Criterion, but these putative clusters do not correspond to geography (**Supplemental Figure 4**). ADMIXTURE calculated a lower cross-validation error for K=2 (0.244) than K=1 (0.260), but again the putative populations do not correspond to geography (**Supplemental Figure 5**). Finally, a genetic distance matrix identified three distinct clusters, but these clusters also do not correspond to geography and are not borne out in a maximum-likelihood phylogeny(**Supplemental Figure 6**).

**Figure 3.**
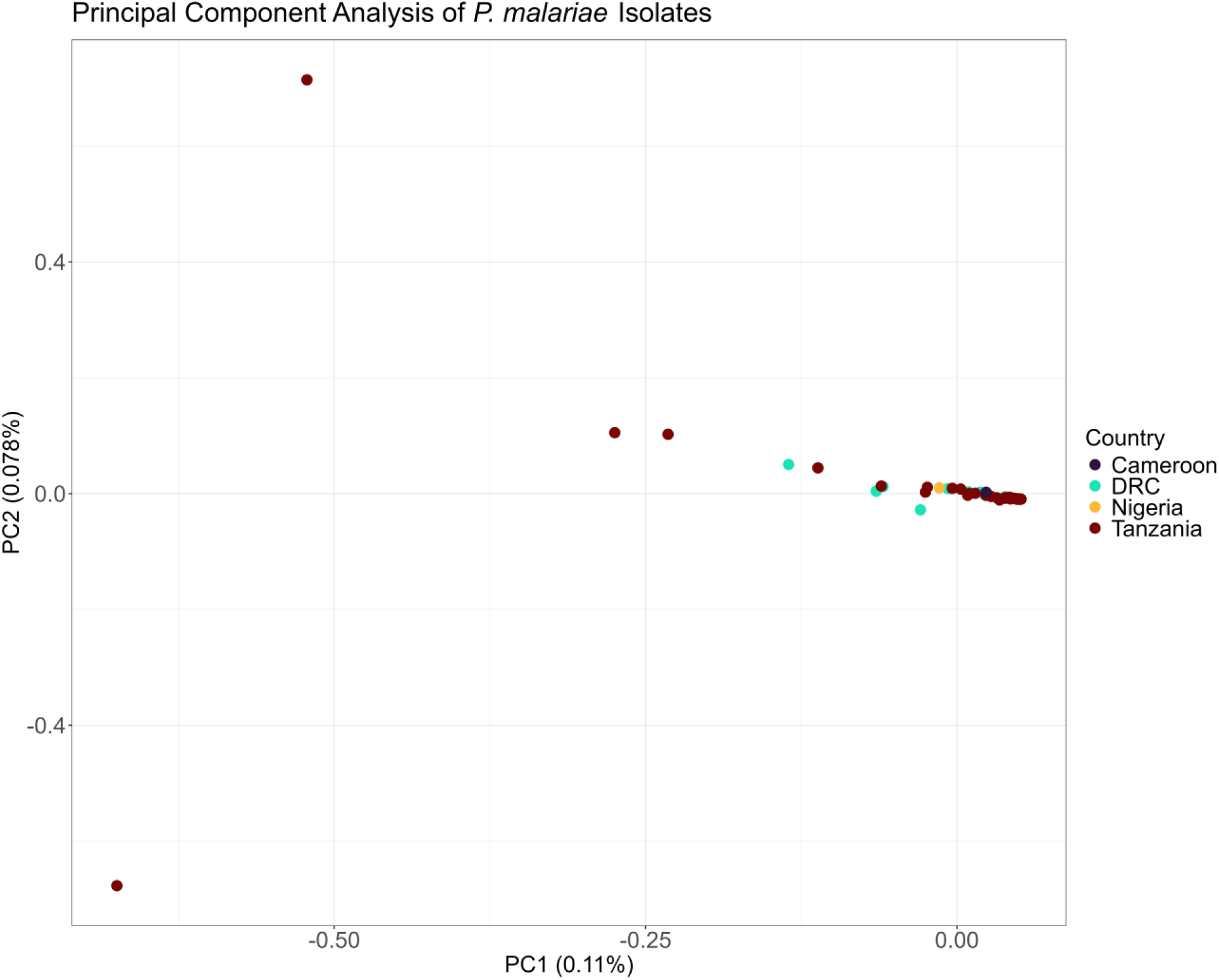
Principal component analysis of monoclonal *P. malariae* isolates. 71 monoclonal *P. malariae* isolates and 178,036 biallelic SNPs are included. The first two principal components (percent of total variation explained) are depicted with isolates colored by country of origin (Cameroon n = 6, DRC n = 16, Nigeria n = 3, Tanzania n = 45).

### *P. malariae* is recovering from a genetic bottleneck

In the absence of compelling evidence for population structure within African *P. malariae* populations, we performed demographic inference modeling to identify the most likely history of this population. Of the five models tested (**Figure 4**, the “three epoch” model, indicating recovery from a bottleneck, was the best fit for African *P. malariae* based on both log likelihood (LL) and the composite-likelihood Akaike Information Criterion (CL-AIC) (**Supplemental Table 1**).

**Figure 4.**
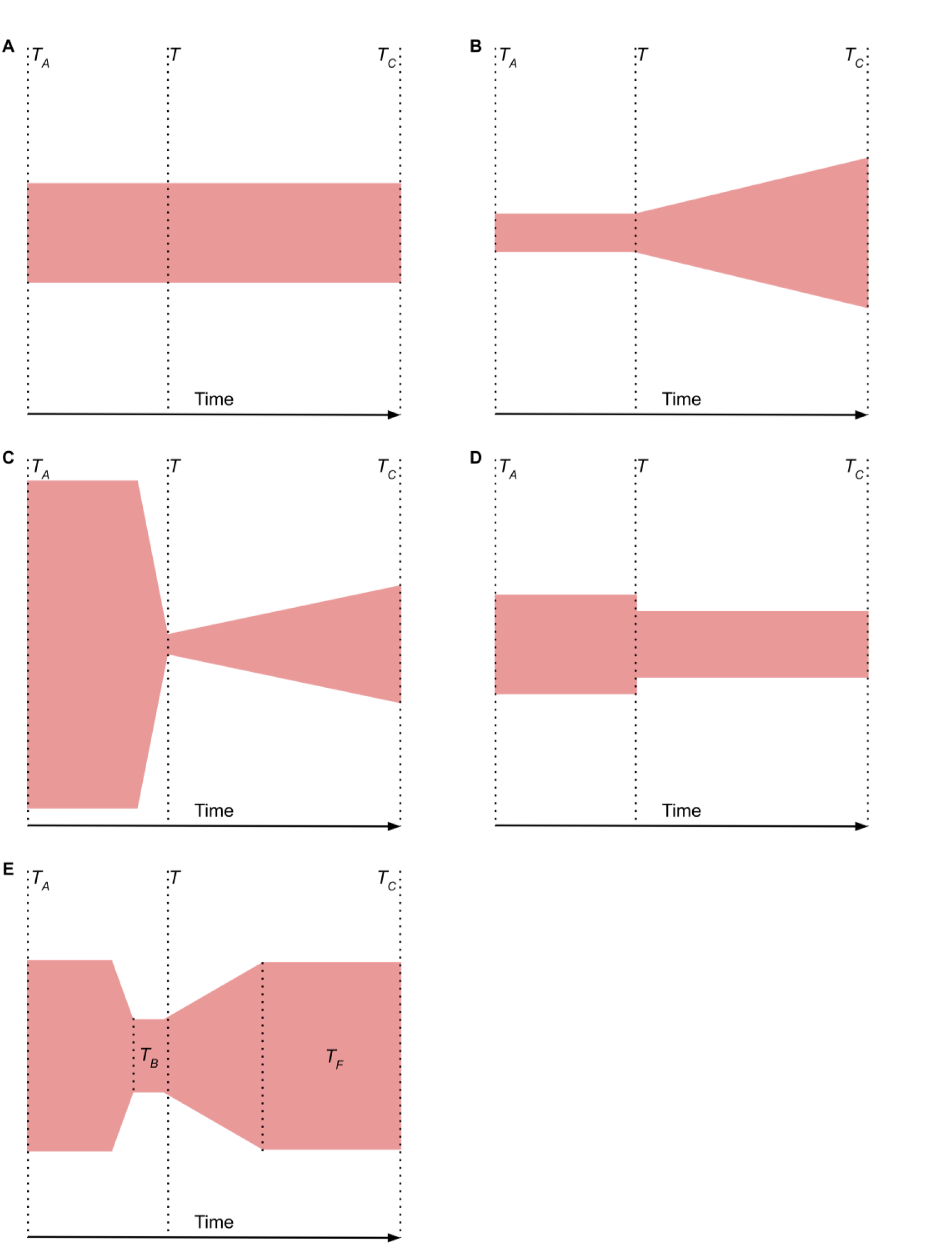
Schematics of demographic models tested for goodness of fit. Plots are organized horizontally, with time in generations on the X-axis and width of colored shape corresponding to effective population size (*N*e). *TA* indicates ancient population while *TC* indicates contemporary population. **A)** Standard neutral model for one population, with no change in population size. **B)** Growth model where population growth begins at time *T*. **C) “**Bottlegrowth” model where an instantaneous size change is followed by exponential time growth at time *T*. **D)** Two epoch model where an instantaneous size change occurs at time *T* followed by a constant *Ne*. **E)** Three epoch model where an instantaneous size change occurs prior to time *T*, with *TB* corresponding to the length of the bottleneck, and bottleneck recovery begins after time *T*, with the time since bottleneck recovery represented by *TF*. Among the five models tested, the three epoch model was the best fit (LL = -514, CL-AIC = -59,663).

### Selective sweeps in orthologs of blood-stage vaccine target and putative antimalarial resistance genes

Genome-wide scans to detect selection identified numerous genes across the genome substantially deviating from neutral expectations. Genome-wide Tajima’s D scans across both genes and exons identified a negative skew (average D = -1.03 and -0.99, respectively), consistent with expectations of population expansion following a bottleneck, or potentially indicating directional selection and/or sweep. However, contrary to our expectations, none of the top hits identified by genome-wide *n*SL or Tajima’s D scans were red blood cell (RBC) invasion ligands or putative antimalarial resistance genes, other than *P. malariae myosin A* (*myoA*)^24^, which has a Tajima’s D value < -2 when scanning across exons, consistent with directional selection (**Supplemental Figure 7, Supplemental Figure 8, Supplemental Table 2, Supplemental Table 3, Supplemental Table 4**).

As such, we adopted a candidate gene approach to scan specific genes of interest due to their putative role in RBC invasion or antimalarial resistance. These genes included apical membrane antigen 1 (*ama1*), chloroquine resistance transporter (*crt*), circumsporozoite protein (*csp*), bifunctional dihydrofolate reductase-thymidylate synthase (*dhfr-ts*), hydroxymethyldihydropterin pyrophosphokinase-dihydropteroate synthase (*pppk-dhps*), Kelch13 (*K13*), liver surface antigen 1 (*lsa1*), multidrug resistance protein 1 (*mdr1*), multidrug resistance protein 2 (*mdr2*), multidrug resistance-associated protein 1 (*mrp1*), multidrug resistance-associated protein 2 (*mrp2*), merozoite surface protein 1 (*msp1*), ookinete surface protein P25 (*P25*), 6-cysteine protein P48/45 (*P48/45*), and thrombospondin-related anonymous protein (*TRAP*). For each of these genes, we performed McDonald-Kreitman (MK) tests to detect evidence of directional selection, as well as all of the tests incorporated in the *DH* software package to identify evidence of sweep.

A significant MK result was found for *lsa1*, but no other genes. *lsa1* also had the second-highest direction of selection, 0.184, which is consistent with weak positive selection (**Table 1**).

**Table 1.**
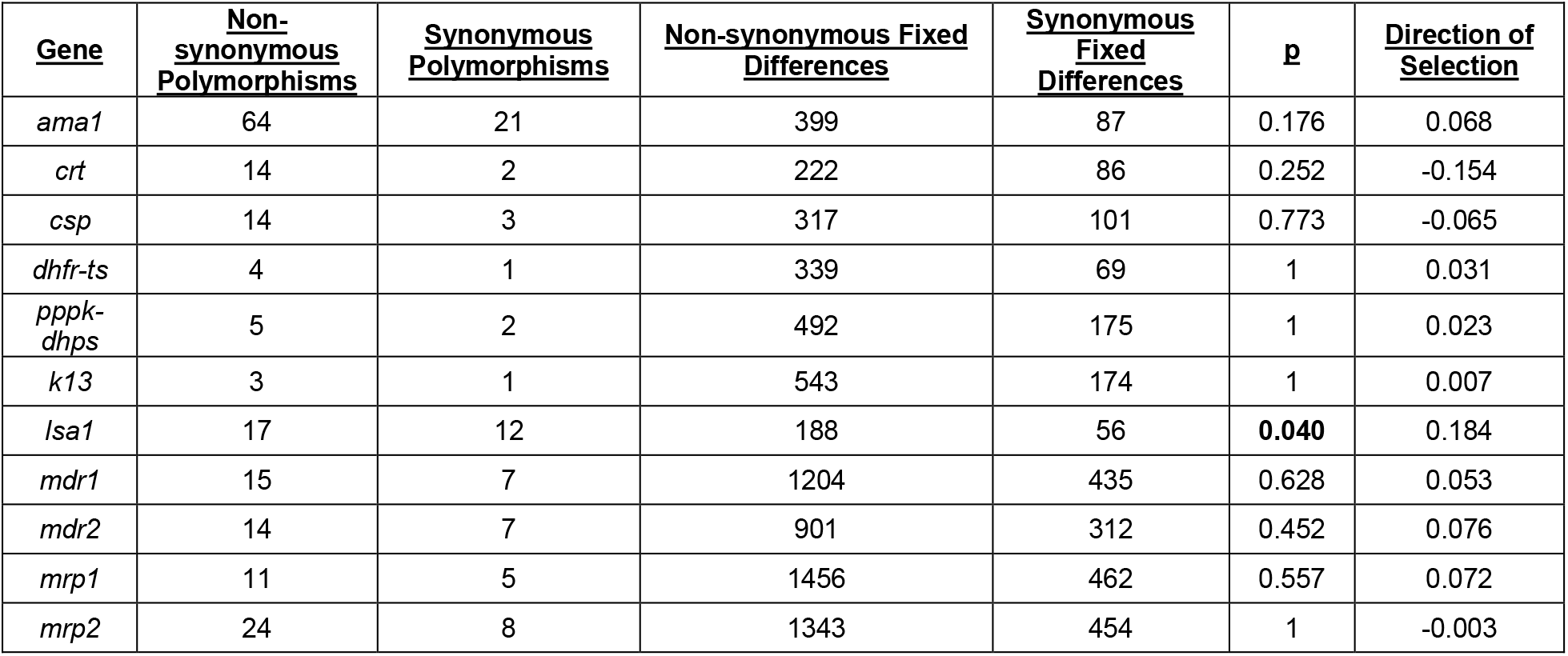
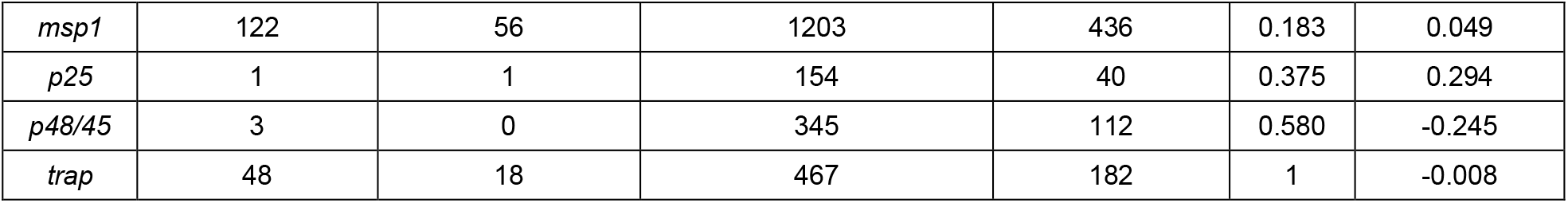
Output of McDonald-Kreitman Tests on Genes of Interest. P values <0.05 are bolded.

| <u>Gene</u> | <u>Non-synonymous Polymorphisms</u> | <u>Synonymous Polymorphisms</u> | <u>Non-synonymous Fixed Differences</u> | <u>Synonymous Fixed Differences</u> | <u>p</u> | <u>Direction of Selection</u> |
| --- | --- | --- | --- | --- | --- | --- |
| *ama1* | 64 | 21 | 399 | 87 | 0.176 | 0.068 |
| *crt* | 14 | 2 | 222 | 86 | 0.252 | -0.154 |
| *csp* | 14 | 3 | 317 | 101 | 0.773 | -0.065 |
| *dhfr-ts* | 4 | 1 | 339 | 69 | 1 | 0.031 |
| *pppk-dhps* | 5 | 2 | 492 | 175 | 1 | 0.023 |
| *k13* | 3 | 1 | 543 | 174 | 1 | 0.007 |
| *lsa1* | 17 | 12 | 188 | 56 | **0.040** | 0.184 |
| *mdr1* | 15 | 7 | 1204 | 435 | 0.628 | 0.053 |
| *mdr2* | 14 | 7 | 901 | 312 | 0.452 | 0.076 |
| *mrp1* | 11 | 5 | 1456 | 462 | 0.557 | 0.072 |
| *mrp2* | 24 | 8 | 1343 | 454 | 1 | -0.003 |
| *msp1* | 122 | 56 | 1203 | 436 | 0.183 | 0.049 |
| *p25* | 1 | 1 | 154 | 40 | 0.375 | 0.294 |
| *p48/45* | 3 | 0 | 345 | 112 | 0.580 | -0.245 |
| *trap* | 48 | 18 | 467 | 182 | 1 | -0.008 |

However, significant D, H, and DH values were identified for six, nine, and ten genes, respectively (**Table 2**). Significant DH p-values consistent with selective sweep were calculated for the RBC invasion genes *ama1, lsa1, msp1*, and *trap*. DH results suggestive of sweep were also found in the putative antimalarial resistance genes *crt, dhfr-ts, pppk-dhps, mdr1, mrp1*, and *mrp2*.

**Table 2.** Output of Tests for Selection on Genes of Interest. P values <0.05 are bolded.

| <u>Gene</u> | <u>Segregating Sites</u> | <u>Tajima's D</u> | <u>p(D)</u> | <u>Fay and Wu's H</u> | <u>p(H)</u> | <u>p(DH)</u> |
| --- | --- | --- | --- | --- | --- | --- |
| *ama1* | 5 | -1.23 | 0.105 | 3.50 | **0.011** | **0.02** |
| *crt* | 4 | -1.82 | **0.005** | -2.20 | **0.031** | **0.003** |
| *csp* | 0 | NA | NA | NA | NA | NA |
| *dhfr-ts* | 2 | -1.106 | 0.176 | -3.05 | **0.021** | **0.007** |
| *pppk-dhps* | 4 | -1.55 | **0.042** | -4.28 | **0.004** | **0.005** |
| *k13* | 0 | NA | NA | NA | NA | NA |
| *lsa1* | 8 | -2.002 | **0.004** | -5.37 | **0.001** | **<0.001** |
| *mdr1* | 4 | -1.75 | **0.012** | -4.44 | **0.003** | **0.001** |
| *mdr2* | 3 | 0.861 | 0.814 | -0.611 | 0.161 | 0.685 |
| *mrp1* | 4 | -1.82 | **0.005** | -4.54 | **0.003** | **<0.001** |
| *mrp2* | 3 | -1.19 | 0.132 | -2.01 | **0.048** | **0.013** |
| *msp1* | 18 | -1.67 | **0.022** | -5.75 | **<0.001** | **0.004** |
| *p25* | 1 | -0.744 | 0.407 | 0.213 | 0.545 | 0.423 |
| *p48/45* | 1 | -0.442 | 0.505 | 0.333 | 0.647 | 0.516 |
| *trap* | 3 | -1.12 | 0.154 | -1.91 | 0.055 | **0.016** |

## Discussion

We present the largest genomic study of *P. malariae* in Africa to-date, enabling a comprehensive analysis of this neglected malaria pathogen in the four high-transmission countries of Cameroon, DRC, Nigeria, and Tanzania. Using hybrid-capture enrichment, we successfully sequenced 77 samples, 71 of which were monoclonal and therefore included in further, more comprehensive analyses. Our study finds that *P. malariae* population genomics differ substantially from those of other human malaria species, including *P. falciparum* isolates from the same countries, with markedly low nucleotide diversity, no geographic population structure, high linkage disequilibrium, and no erythrocyte invasion ligands or antimalarial resistance gene orthologs identified in genome-wide selection scans. This suggests *P. malariae* may rely on markedly different strategies to persist in human populations, perhaps related to its ability to persist in human hosts undetected for months to years.

As has been seen in other non-falciparum species, COI was significantly lower in *P. malariae* than in *P. falciparum* isolates from the same countries. This trend was also observed in African *P. ovale* spp. infections^25^, and may reflect the lower prevalence and lighter transmission intensity of these non-falciparum species. In *P. malariae*, low COI may also correspond to its low genetic diversity and/or frequent transmission of the same clones. It is also notable that *P. malariae* has most often been detected as part of mixed infections with *P. falciparum*^7–10^.

In contrast to *P. ovale curtisi*^25^ and *P. vivax*^26^, *P. malariae* exhibits significantly lower nucleotide diversity than *P. falciparum* when comparing 1-1 orthologs. While a similar trend is also observed in *P. ovale wallikeri*^25^, the magnitude of the difference in *P. malariae* is much greater. The extremely low nucleotide diversity within *P. malariae* is likely a relic of the relatively recent bottleneck that occurred following zoonosis from the non-human primates where *P. malariae* originated^17^ and corresponds to a low effective population size (*Ne*). The lingering impacts of this bottleneck and corresponding low *Ne* are also reflected in the apparent lack of geographic population structure within African *P. malariae*. That lack of geographic population structure is supported by the failure of multiple methods to identify a consistent and parsimonious trend, including PCA, DAPC, ADMIXTURE, and maximum likelihood phylogenetics. Further support comes from the high linkage disequilibrium observed across the *P. malariae* genome; this delayed decay of LD over distances in the *P. malariae* genome suggests fewer recombination events between diverse strains over time.

While our analyses provide strong support for *P. malariae* experiencing a recent bottleneck, they also indicate that it is currently in recovery. The average genome-wide Tajima’s D value is negative, which is consistent with recovery from a bottleneck and/or selective sweep. We find significant evidence of selective sweep in genes of interest, as well as strong evidence from demographic inference modeling that *P. malariae* is in the recovery phase, with the three-epoch model having the best fit. Specifically, within *P. malariae*, that model could imply 1) an ancient population of non-human primate parasites that 2) experienced a severe bottleneck due to spilling over to human hosts and finally 3) began to recover from that bottleneck. While the African *P. malariae* population is still low in diversity and lacking in population structure, the model and our results suggest that *Ne* is on an upward trajectory.

*P. malariae* is highly unusual among human malaria parasites in that genome-wide selection scans identified neither erythrocyte invasion ligands nor antimalarial resistance orthologs as sites of notable balancing or directional selection using Tajima’s D and *n*SL. This difference is particularly notable in contrast with African *P. ovale* spp. from some of the same study regions, where multiple such genes had some of the highest values for both statistics^25^. However, the candidate gene approach detected signatures of selection in four erythrocyte invasion ligands (*pmama1, pmlsa1, pmmsp1*, and *pmtrap*) and six antimalarial resistance orthologs (*pmcrt, pmdhfr-ts, pmpppk-dhps, pmmdr1, pmmrp1*, and *pmmrp2*). The lack of identification of these genes in genome-wide scans may be due to the bottleneck recovery, along with the remarkably high LD in *P. malariae*.

Of the genes analyzed with the candidate gene approach, only *pmlsa1* shows evidence of positive selection. This trend is unusual given that erythrocyte invasion ligands in other human *Plasmodium* spp. typically show evidence of balancing selection^25,27,28^. This finding may, however, reflect the susceptibility of the McDonald-Kreitman test to bias from demographic factors and slightly deleterious mutations^29,30^. Nonetheless, the signatures of selective sweep in *pmlsa1*, as well as signatures of positive selection at four erythrocyte invasion ligands point to a consistent trend within *P. malariae*. This sweep may have arisen as part of the process of adapting to human hosts following the zoonotic event, in which case mutations in either these genes or those with which they are linked would likely have provided a selective advantage.However, it is also possible that the sweep stems from the bottleneck itself, particularly if there has been more than one severe bottleneck^31^.

Selective sweep in the antimalarial resistance orthologs is more precedented^25,32–34^ within other *Plasmodium* spp. Given that *P. malariae* is most commonly found in mixed-species infections, it is subject to drug pressure like *P. falciparum*. However, as with the erythrocyte invasion ligands, it is also possible that sweep in these antimalarial resistance orthologs is more related to bottleneck recovery than to drug pressure.

This study is subject to several limitations. As with all *Plasmodium* genomic studies, our ability to perform robust enrichment and high-coverage sequencing was limited by parasitemia, meaning that we could only thoroughly analyze relatively high-density infections. However, the hybrid capture enrichment method is able to enrich much lower density infections than direct sequencing. Second, sampling for the parent studies was not nationally representative. While we have samples spanning 14 regions of mainland Tanzania, the geographic spread of the other three countries in this analysis is restricted to specific regions. While it is possible that we have failed to detect population structure that would only be apparent with the inclusion of samples from other regions, such as West Africa, the Horn of Africa, and Southern Africa, the observed lack of population differentiation between such disparate locations as coastal Tanzania and Nigeria suggests that such structure would be weakly geographically correlated if present at all. In addition, a previous microsatellite study that includes isolates from West Africa found the same lack of geographic separation^19^. Third, the incomplete nature of the available *P. malariae* genome assembly used both to design our RNA baits and to align sequencing reads, may have led us to miss variants and trends in biologically important regions of the genome which are currently not assembled in chromosomes. Fourth, the study design used for collection of these isolates varied, which may have introduced unexpected biases. Finally, the comparatively low positivity rates for *P. malariae* as compared to *P. falciparum* inherently limit which samples can be used for analysis. Nevertheless, this study remains the largest conducted to-date and enables genomic analyses across multiple geographies and populations.

This study augments our understanding of the population genomics and demographic history of the largely neglected malaria parasite, *P. malariae*, in Africa. As has been suggested by other studies, we find that *P. malariae* is a genomically atypical human malaria parasite, likely owing to the impact of a zoonotic spillover event and associated genetic bottleneck in its past. The lack of geographic population structure within Africa and the extremely low nucleotide diversity present unique challenges for genomic surveillance and molecular epidemiology, as they are likely to complicate efforts to track importation and transmission networks. While *P. malariae* is currently a relatively minor problem in Africa, there is evidence that this may change in the future^13,35–38^. As such, it may become desirable to tailor national drug strategies and/or design vaccines to target *P. malariae* as well as *P. falciparum*, in addition to monitoring the impact of interventions targeting *P. falciparum* on the *P. malariae* population. Such efforts will need to contend with the selective sweeps identified in many of the relevant loci. While there are limited studies of *P. malariae* genomics within Africa, there are even fewer elsewhere. Evidence of population differentiation between African and Thai isolates suggest that other malaria-endemic regions may not necessarily exhibit the same trends seen in this analysis. As such, epidemiological as well as genomic studies in Asia and the Americas are warranted.

## Methods

DNA was isolated from 16,596 blood samples from six malaria studies conducted across sub-Saharan Africa using standard Chelex methods^39^. Samples were either dried blood spots or whole blood samples leukodepleted at the time of collection by CF11 filtration^40^. DNA isolates were screened by real-time polymerase chain reaction for amplification of the *P. malariae* 18S rRNA gene (**Table 3**), as described elsewhere^11^. Positive samples were considered for sequencing based on geographic diversity and amplification of the 18S rRNA gene before 36 cycles.

**Table 3.** Details of sample collection for samples included in study.

| <u>Study</u> | <u>Country</u> | <u>Year</u> | <u>Population</u> | <u>Number Screened</u> | <u>Number positive for Pm</u> | <u>Number included</u> |
| --- | --- | --- | --- | --- | --- | --- |
| Molecular Surveillance of Malaria in Tanzania (MSMT)<sup>9,10</sup> | Tanzania | 2021 | Symptomatic individuals reporting to health facilities in 10 regions, asymptomatic individuals in three regions | 4,712 (3284 symptomatic, 1428 asymptomatic) | 72 (55 symptomatic, 17 asymptomatic) | 39 (34 symptomatic, 5 asymptomatic) |
| Group Antenatal Care (GANC)<sup>41,42</sup> | Tanzania | 2021 | children under five in Geita region | 734 | 5 | 3 |
| Transmission from Submicroscopic Malaria in Tanzania (TranSMIT)<sup>43,44</sup> | Tanzania | 2018-22 | Asymptomatic individuals in schools and health clinics in rural Bagamoyo district | 1964 | 66 | 31 |
| Kinshasa Malaria Longitudinal Cohort<sup>7</sup> | DRC | 2015-17 | Members of households participating in longitudinal malaria study | 9089 | 321 | 16 |
| Genomic Epidemiology of *P. malariae* in Southern Nigeria<sup>45</sup> | Nigeria | 2017-18 | Hospital and community survey | 58 | 7 | 4 |
| Pan-African Genomic Epidemiology Network (PAMGEN) | Cameroon | 2021 | Pregnant women at antenatal visit | 39 | 8 | 7 |

### Library preparation and sequencing

Selected DNA isolates were fragmented enzymatically and underwent library preparation using the Twist Library Preparation EF 2.0 kit (Twist Bioscience, South San Francisco, CA). Libraries then underwent parasite DNA enrichment using a custom-designed Twist hybridization capture protocol using 280,042 RNA baits specifically designed to amplify *P. malariae* genomic DNA across the full genome without reacting with background human DNA. Enriched samples were then amplified, purified, and submitted for Illumina short-read 150bp sequencing on the NovaSeq 6000 S4-XP system with paired-end chemistry.

### Sequencing data alignment and variant calling

*fastqc* v0.12.1 (https://www.bioinformatics.babraham.ac.uk/projects/fastqc/) was used to check the quality of raw sequencing reads before trimming sequencing adapters using *Trim Galore* v0.6.7 (https://www.bioinformatics.babraham.ac.uk/projects/trim_galore/). Trimmed reads were then competitively aligned to the *P. malariae* (PmUG01 strain), *P. falciparum* (Pf3D7), and *Homo sapiens* (Hg38 strain) using BBSplit within BBMap v38.96 (https://sourceforge.net/projects/bbmap/). Reads that best aligned to the *P. falciparum* and *H. sapiens* genomes were discarded, and the remaining reads were then aligned to the PmUG01 reference genome with *bwa-mem2* v2.2.1^46^. *picard* v2.26.11 (https://broadinstitute.github.io/picard/) was then used to sort and deduplicate reads before variant calling with GATK v4.5.0.0^47^. Coverage statistics were calculated using *samtools* v1.20^48,49^.

Variants were called across each sample following the *GATK* best practices pipeline^47^. Variants were called using gVCF mode of the Haplotype Caller function within each chromosome of each individual sample before calling variants across all samples^50^. SNPs were then filtered with GATK if they met the following thresholds: quality by depth <2.5, Fisher strand bias >10, mapping quality >50, mapping quality rank sum <-2.5, read position rank sum <-2.5. The resulting filtered VCF was then filtered to biallelic SNPs using bcftools v1.2.0^49^ and SNPs with greater than 20% missingness were excluded with vcftools v0.1.15^51^. SNPs falling within the hypervariable *PIRs* and SNPs within tandem repeats as identified by *Tandem Repeats Finder* v4.09.1^52^ were masked in vcftools. Finally, only SNPs within the 14 chromosomes were retained for further analysis, with all SNPs on the mitochondrion, apicoplast, and extrachromosmal contigs excluded with bcftools.

### Selection of matched *P. falciparum* samples

For comparison with sympatric *Plasmodium falciparum* parasites, whole-genome sequencing data from 384 *P. falciparum* isolates from the same countries of origin as the *P. malariae* isolates (121 from Cameroon, 93 from DRC, and 170 from Tanzania -while Nigerian data were available, the Cameroonian data were more geographically proximate to the Nigerian *P. malariae* isolates) were selected from the publicly-available *Pf7* dataset^22^.

### Nucleotide diversity

The *OrthoMCL*^53^ database implemented in *PlasmoDB*^54^ was used to identify orthologous protein-coding genes between the *P. malariae* and *P. falciparum*, and 3,763 one-to-one orthologs were analyzed to compare nucleotide diversity between the two species. Orthologs were masked based on the masking steps outlined above and also excluded if they were found outside of the *P. falciparum* core genome. In addition, ortholog pairs were excluded unless 60% of samples of each species had ≥ 5X coverage of the gene at least every 10 base pairs. Of the 3,763 candidate orthologs, 1,377 unmasked ortholog pairs were retained for final analyses. Nucleotide diversity (π) was then calculated in vcftools over each ortholog in each species, and the distribution of π by species was compared with a Student’s t-test.

### Linkage disequilibrium and complexity of infection

Linkage disequilibrium across genomic intervals in each species was calculated using *PLINK* v1.90b7.2^23^ with 100 bp windows and a minor allele frequency cutoff of 0.01. Complexity of infection was estimated for each isolate using *coiaf* v0.1.2^21^ with a minor allele frequency cutoff of 0.05. Data visualizations were constructed in *R* v4.2.2^55^.

### Population Structure and Demography

*vcfdo* (https://github.com/IDEELResearch/vcfdo) was used to calculate within sample allele frequency (WSAF) among monoclonal samples, then filtered to SNPs with WSAF equal to 0 or 1, as heterozygous calls in monoclonal haploid infections are likely to represent sequencing errors or paralogous misalignments. Principal component analysis (PCA) of monoclonal isolates was performed in *PLINK*. Population structure among monoclonal isolates was also assessed using *ADMIXTURE* v1.3.0^56^, discriminant PCA in *adegenet* v2.1.10^57,58^, genetic distance in *fastreeR* (https://github.com/gkanogiannis/fastreeR), and maximum likelihood phylogeny in *RAxML Next Generation* v1.2.2 (https://github.com/amkozlov/raxml-ng) following format conversion using *vcf2phylip* v2.8 (https://github.com/edgardomortiz/vcf2phylip).Demographic history was initially inferred using *donni*^59^ to determine rough estimates for one-population model parameters using the following models implemented in ∂a∂i^60^: “bottlegrowth_1d”, “growth”, “snm_1d”, “two_epoch”, and “three_epoch”. The 95% confidence intervals (CI) from the *donni* output were then used as bounds for optimization in *dadi-cli*^61^.Because of the high linkage in *P. malariae*^62^, model fit was compared using the composite likelihood Akaike Information Criterion (CL-AIC)^63,64^, calculated in a custom R script. To perform this calculation, the Godambe Information Matrix^62^ script in *dadi-cli* was modified to extract the *H* and *J* matrices for the 95% CI with 0.01 step size. All converged optimizations from the best fit file for each model were then used to determine median log likelihood values and median demographic parameter estimates.

### Signatures of Selection

Among monoclonal isolates of each species, *n*SL^65^ was calculated at each SNP using *selscan* v2.2.0^66^ and the SNPs with absolute values in the top 0.5% were intersected with the PmUG01 genome annotation using the *ape* v5.8^67^, *GenomicRanges* v1.50.2^68^, and *plyranges* v1.18.0^69^ Rpackages. Tajima’s D was calculated in windows of 300 bp with a 10bp step size across the entire genome with *vcftools*; the top 0.5% (absolute value) of windows were analyzed as above.

To detect signs of selection within blood-stage vaccine target orthologs and antimalarial resistance gene orthologs (*ama1, crt, csp, dhfr-ts, Kelch13, lsa1, mdr1, mdr2, mrp1, mrp2, msp1, p25, p48-45, pppk-dhps, trap*), *bcftools* was used to generate fasta sequences for both coding sequences (CDS) and full gene sequences for each gene within each isolate for both *P. malariae* and *P. falciparum*, then sequences were aligned using MAFFT v7.490^70^. The CDS alignments were imported into DnaSP v6.12.03^71^, where the McDonald-Kreitman^72^ test for positive selection was calculated. Because the neutrality index is subject to bias, we calculated and reported direction of selection (DoS) instead^73^. The full sequence alignments of *P. malariae* sequences with one randomly selected *P. falciparum* outgroup sequence were analyzed with the *Readms* module of *DH* (https://github.com/drkaizeng/publications-and-software/blob/main/dh/dh.zip), which calculates Tajima’s D^74^, Fay and Wu’s H^75^, a p-value for the DH test^76^, and the E test^76^ over 10,000 coalescent simulations. The DH test is unique in its sensitivity to selective sweeps and insensitivity to demographic forces, whereas Tajima’s D and Fay and Wu’s H are both prone to demographic bias^76^.

## Data Availability

Parasite sequence data is available through SRA (BioProject ID PRJNA1157442).

## Data availability

Parasite sequence data is available through SRA (BioProject ID PRJNA1157442).

## Code availability

Code used for analysis is available from GitHub at https://github.com/IDEELResearch/PmPopGen

## Acknowledgements

Kyaw Lay Thwai, Claudia Gaither, Meredith Muller, and Srijana Chhetri provided laboratory assistance at UNC. Andrew Guinness wrote the R function to calculate CL-AIC. Professor Martin Meremikwu (University of Calabar Teaching Hospital) and Dr. Dr Tobias Apinjoh (University of Buea) provided invaluable support for field collections in Nigeria and Cameroon, respectively. The authors wish to thank participants and parents or guardians of all children who took part in the studies which were undertaken. We appreciate the support of the in-country teams, and other colleagues including community and health facility staff who took part in data collection or laboratory processing of samples.

## Ethics declarations

As part of the parent studies from which samples were derived, written informed consent, assent, and/or parental consent was obtained from all participants.Institutional review board approvals were obtained for parent studies as follows: Cameroon, from the Gambia Government/MRCG Joint Ethics Committee (REF SCC1626); DRC, from the University of North Carolina at Chapel Hill (IRB#: 14-0489) and the Kinshasa School of Public Health (ESP/CE/015/014); Nigeria, from the Cross River State Health Research Ethics Committee (REC No CRSMOH/RP/REC/2017/809). For the MSMT samples, the study protocol was submitted to the Tanzanian Medical Research Coordinating Committee (MRCC) of the National Institute for Medical Research (NIMR) for review and ethical approval, which was granted. The protocol was also submitted for review and approval by the ethics committee of WHO in Geneva, Switzerland, which was also granted. All research participants were asked and provided individual consent (or assent for children aged 7–17 years of age) for their participation in the survey and biobanking for future research. For children under the legal age of adulthood in Tanzania (< 18 years), consent was obtained from a parent or guardian. An informed consent form was developed in English and translated in Kiswahili and used to obtain consent both verbally and in writing from all participants. All participants agreed and signed the consent or assent form or provided a thumbprint in conjunction with the signature of an independent witness in case the study participant was illiterate. All experiments were performed in accordance with relevant guidelines and regulations in accordance with the Declaration of Helsinki. For the GANC samples, the study protocol was approved by the National Health Research Ethics Sub-Committee (NatHREC) of the Ministry of Health, Community Development, Gender, Elderly and Children (Dar es Salaam, Tanzania) and the US Centers for Disease Control and Prevention (CDC; Atlanta, GA, USA) institutional review board.

