## Supplemental Material for "Population Genomics of *Plasmodium malariae* from Four African Countries"

**Table of Contents**

Figure S1………………………………………………………………………………………………….3

Figure S2………………………………………………………………………………………………….4

Figure S3………………………………………………………………………………………………….5

Figure S4………………………………………………………………………………………………….6

Figure S5………………………………………………………………………………………………….7

Figure S6………………………………………………………………………………………………….8

Figure S7………………………………………………………………………………………………….9

Figure S8…………………………………………………………………………………………………10

Table S1………………………………………………………………………………………………….11

Table S2………………………………………………………………………………………………….12

Table S3………………………………………………………………………………………………….20
Table S4………………………………………………………………………………………………….24

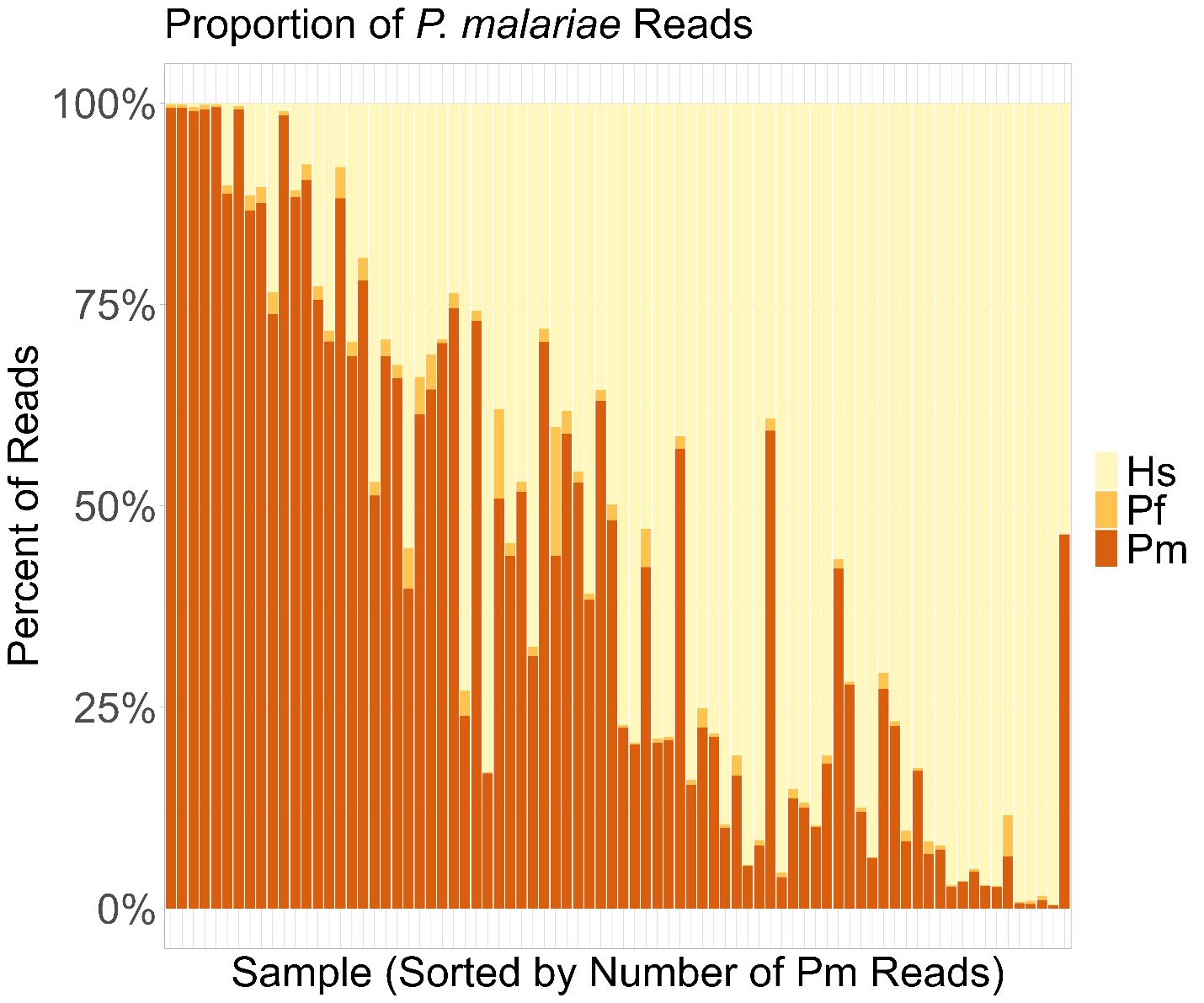

**Supplemental Figure 1 – Enrichment of P. malariae reads by sample.** Percent of sequencing reads aligning to Homo sapiens (Hs), P. falciparum, and P. malariae genomes for 81 samples.

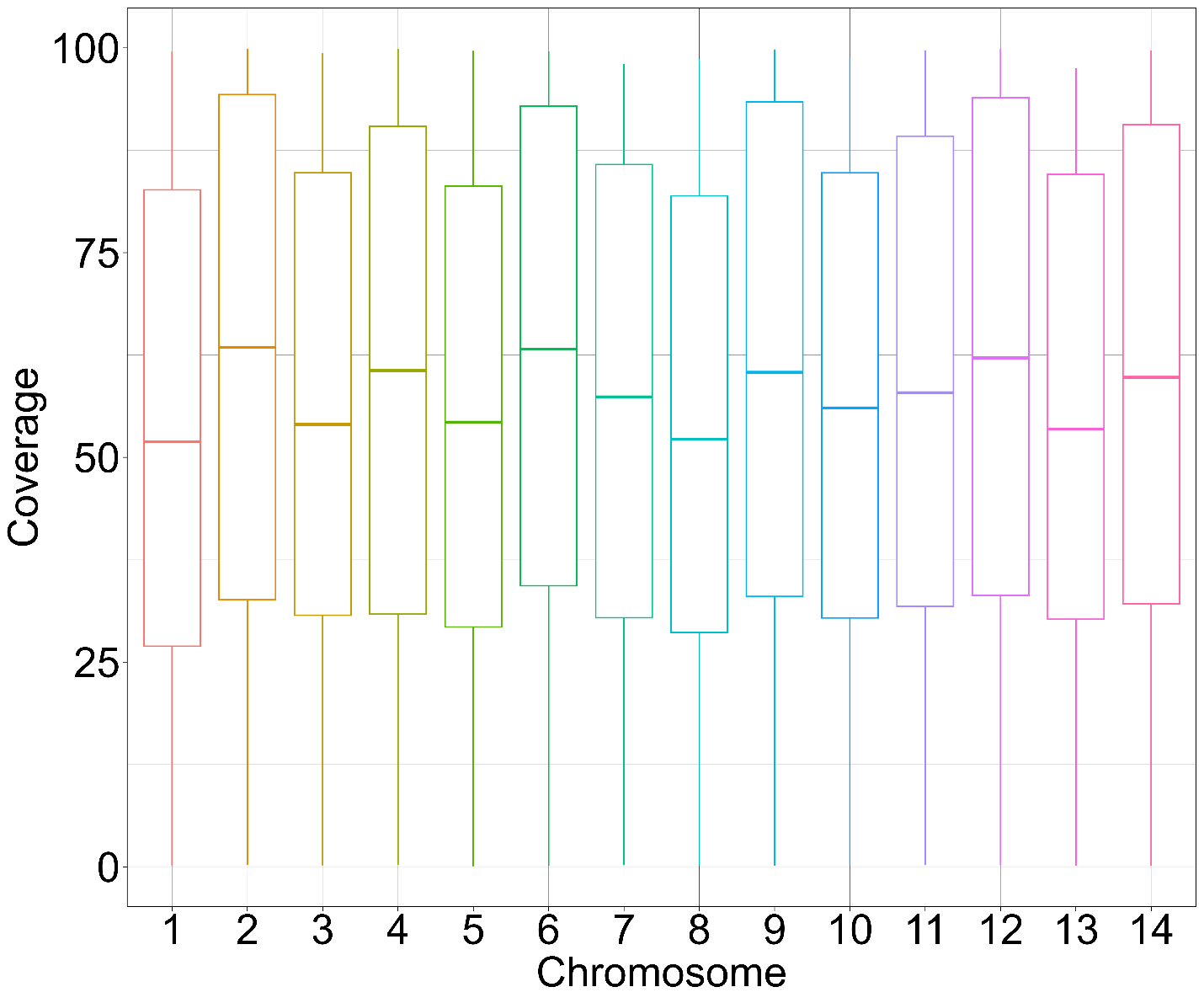

**Supplemental Figure 2 – Coverage by chromosome across all samples.** Boxes highlight 25^th^, 50^th^, and 75^th^ percentiles, showing mean values above 50X coverage across all 14 chromosomes.

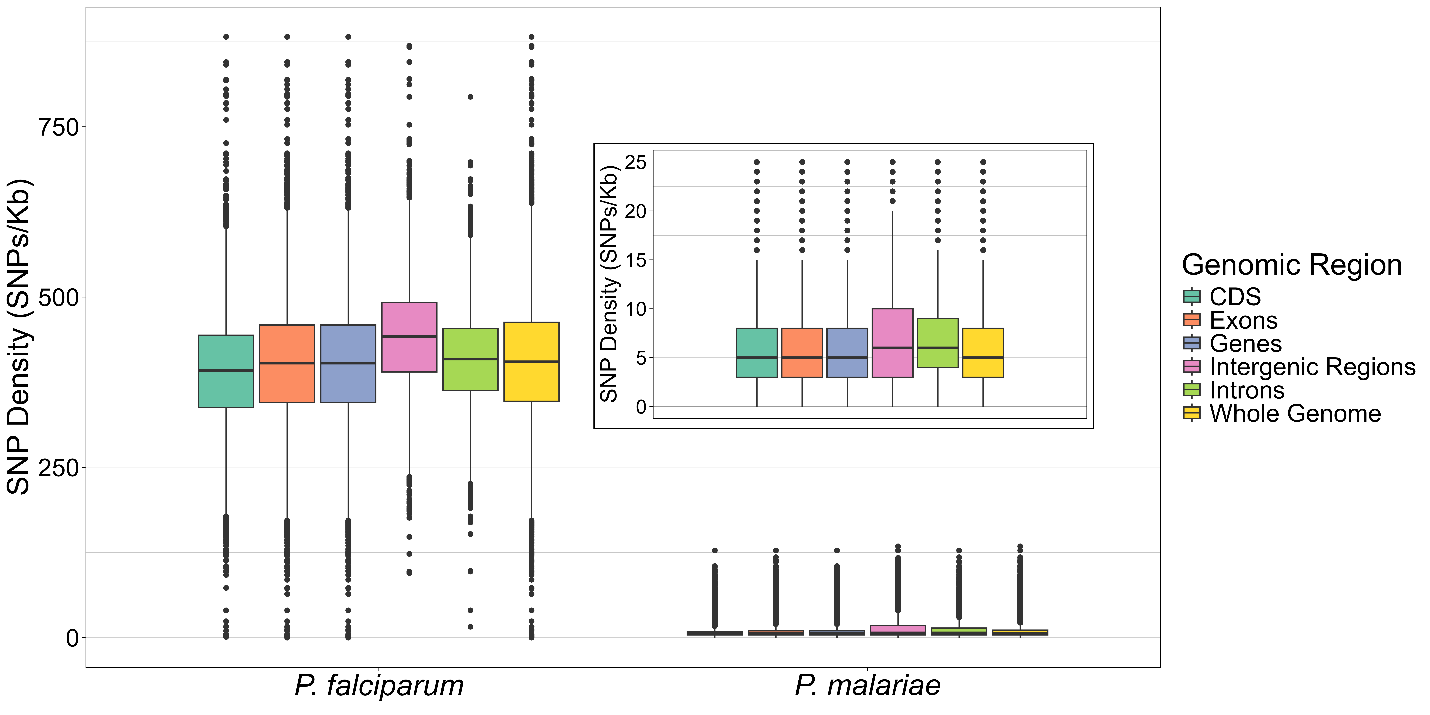

**Supplemental Figure 3 – SNP density across different genomic regions in P. falciparum and P. malariae.** Inset plot shows P. malariae values with smaller scale. Boxes highlight 25^th^, 50^th^, and 75^th^ percentiles. While no statistical analysis was performed, SNP density is clearly lower in P. malariae than in P. falciparum.

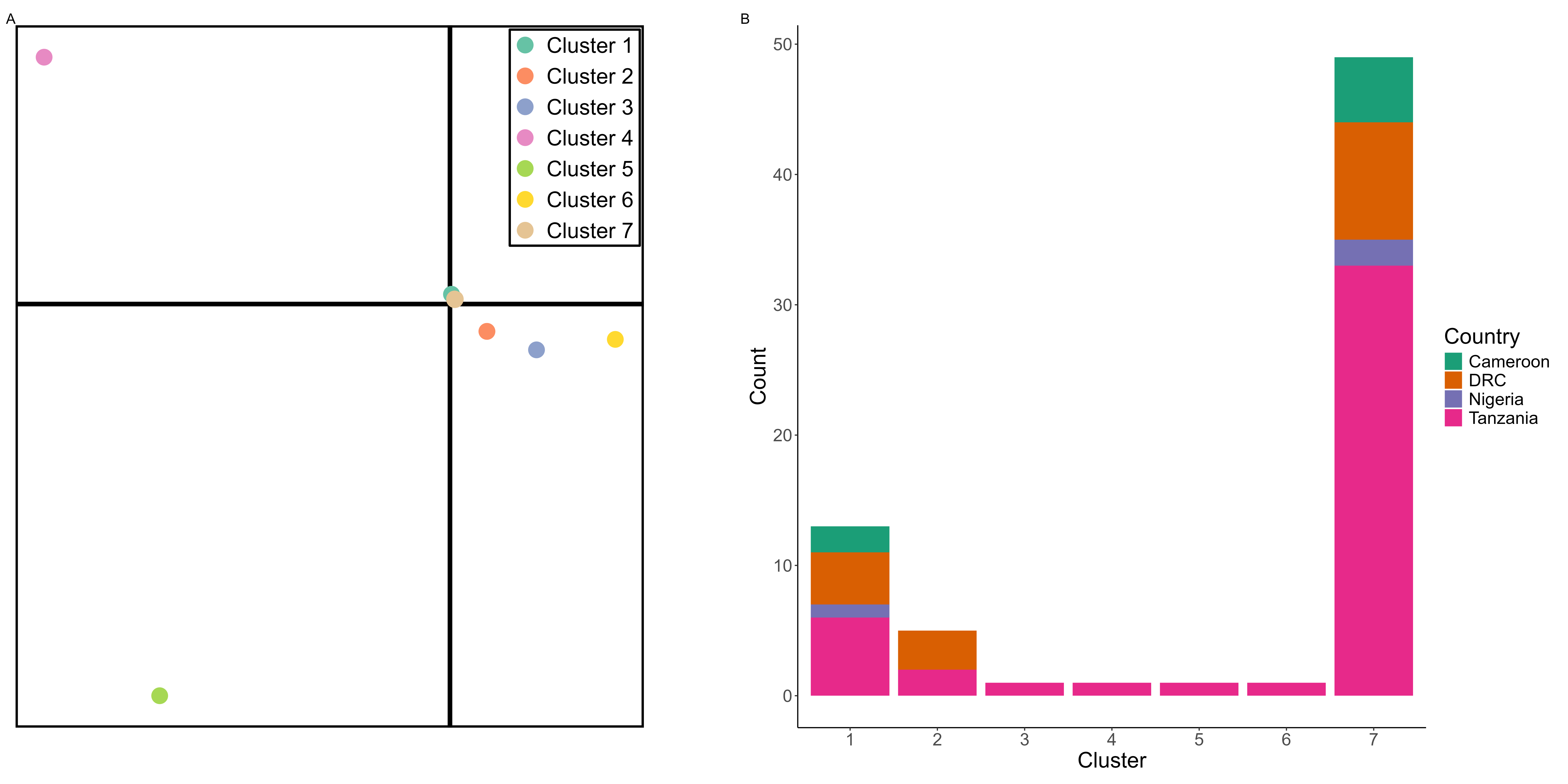

**Supplemental Figure 4 – A) Discriminant PCA showing seven putative population clusters within 71 monoclonal African P. malariae samples. B) Stacked bar plot of samples belonging to each population cluster by country of origin**. No geographic separation is visible.

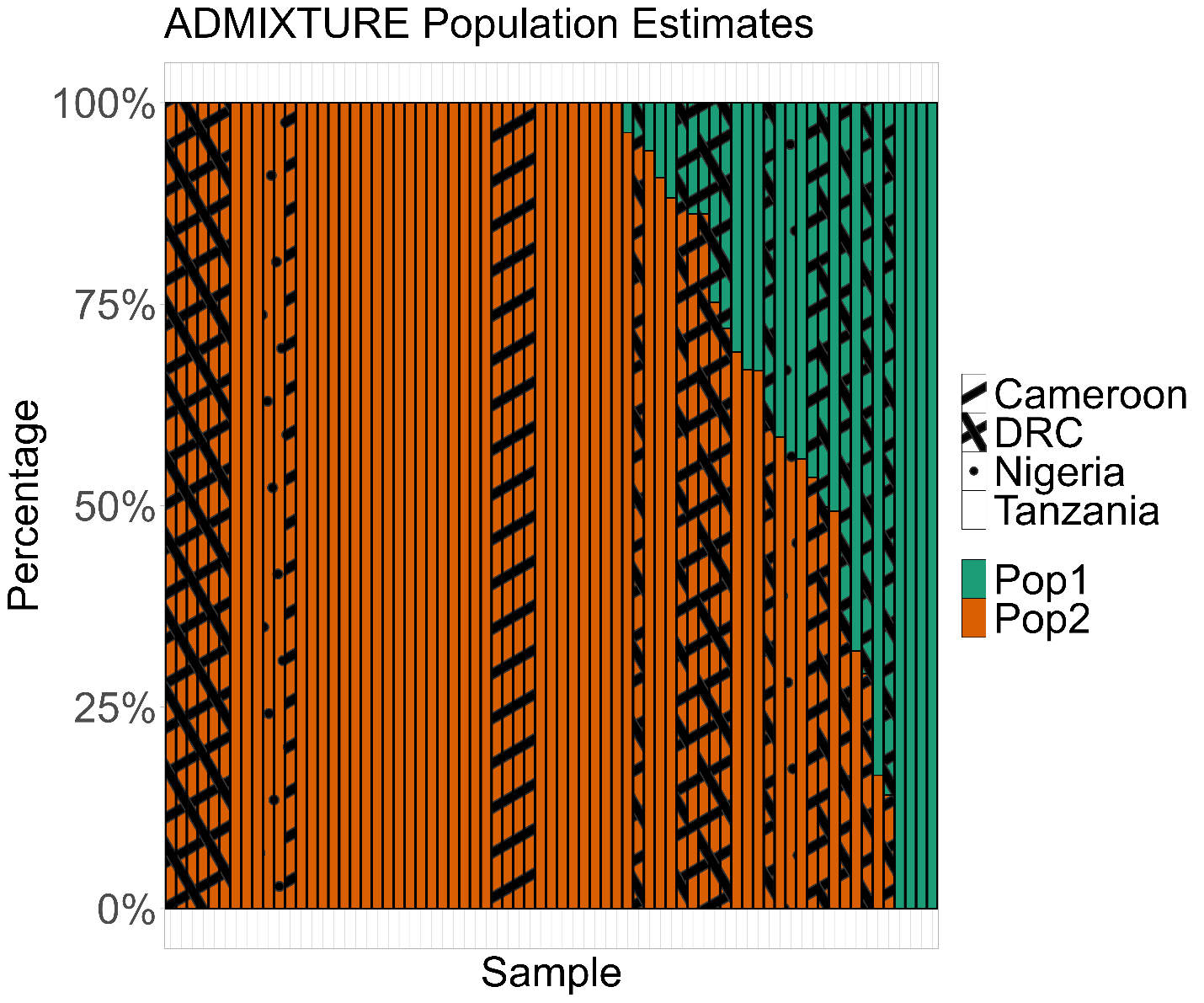

**Supplemental Figure 5 – ADMIXTURE plot of population identity for 71 monoclonal P. malariae isolates with K=2 (the value with the lowest cross-validation error)**. Fill pattern indicates geographic origin. While all Cameroon samples cluster together within putative population 2, no other clear geographic separation is visible.

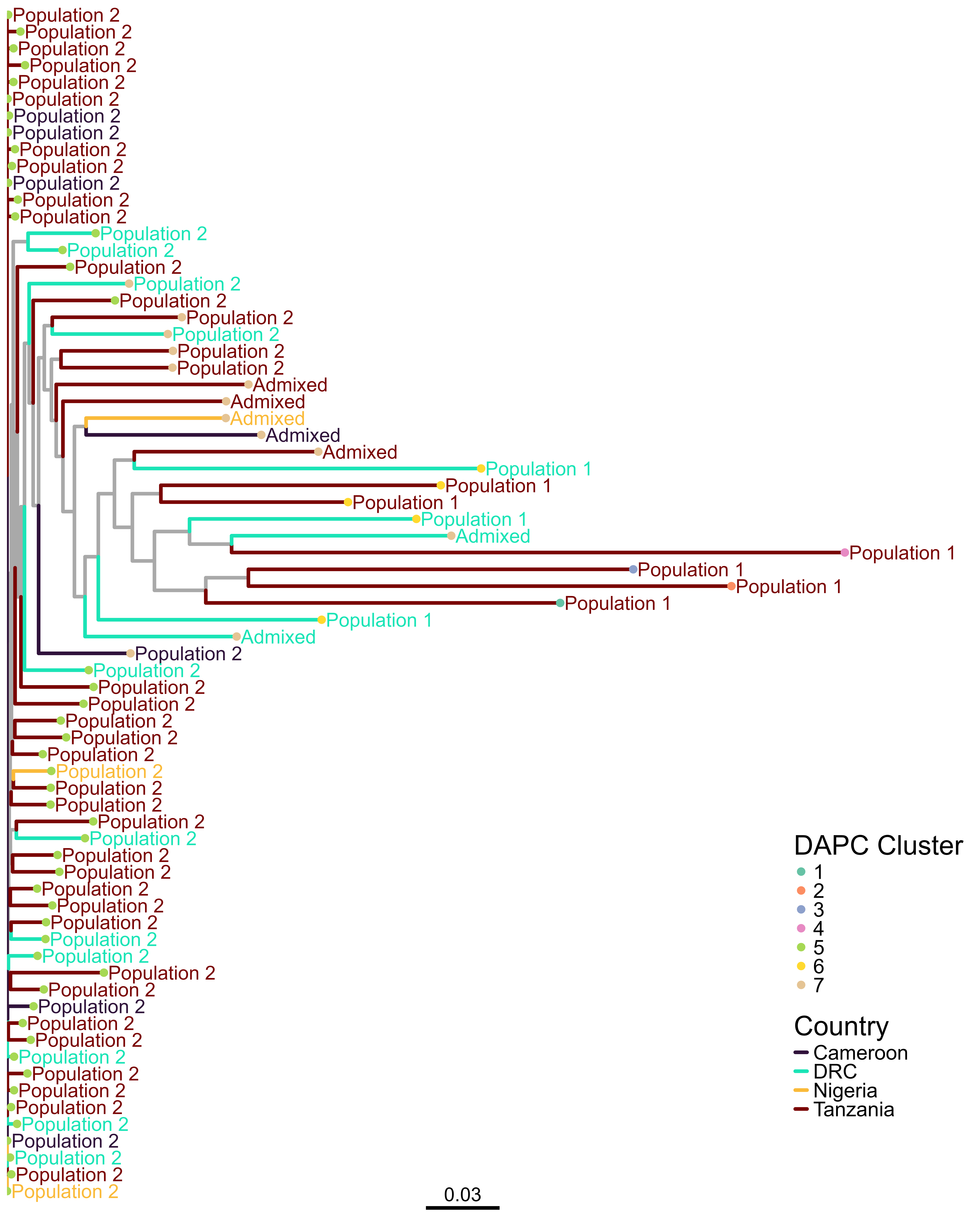

**Supplemental Figure 6 – Maximum likelihood phylogeny of 71 monoclonal P. malariae isolates.** Phylogeny was generated with RAxML Next Generation, using 10 starting trees and 50 bootstraps. Branches are colored by country of origin with grey used to indicate interior nodes that do not reflect known isolates. Text labels correspond to the primary population determined by ADMIXTURE (see **Supplemental Figure 5**). Samples with population proportions between 40% and 60% are considered to be admixed. Finally, tip label colors correspond to DAPC clusters (see **Supplemental Figure 4**).

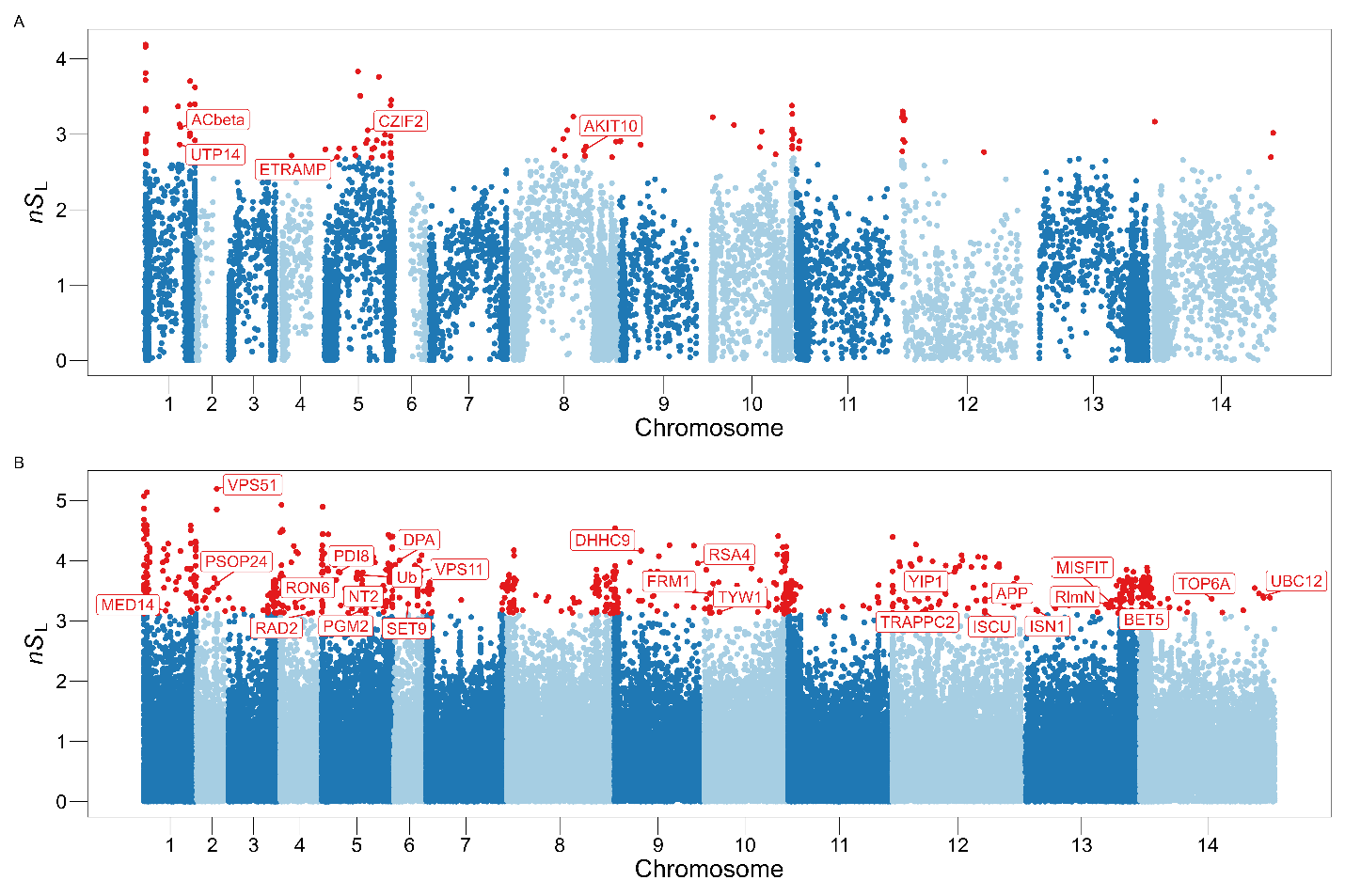

**Supplemental Figure 7 – Genome-wide nS_L_ values with A) a minor allele frequency cutoff of 0.05 applied and B) no minor allele frequency cutoff applied**. Points in red are in the top 0.5% of absolute nS_L_ values. Annotated genes are labeled, none of which are blood-stage vaccine candidate orthologs or putative antimalarial resistance genes.

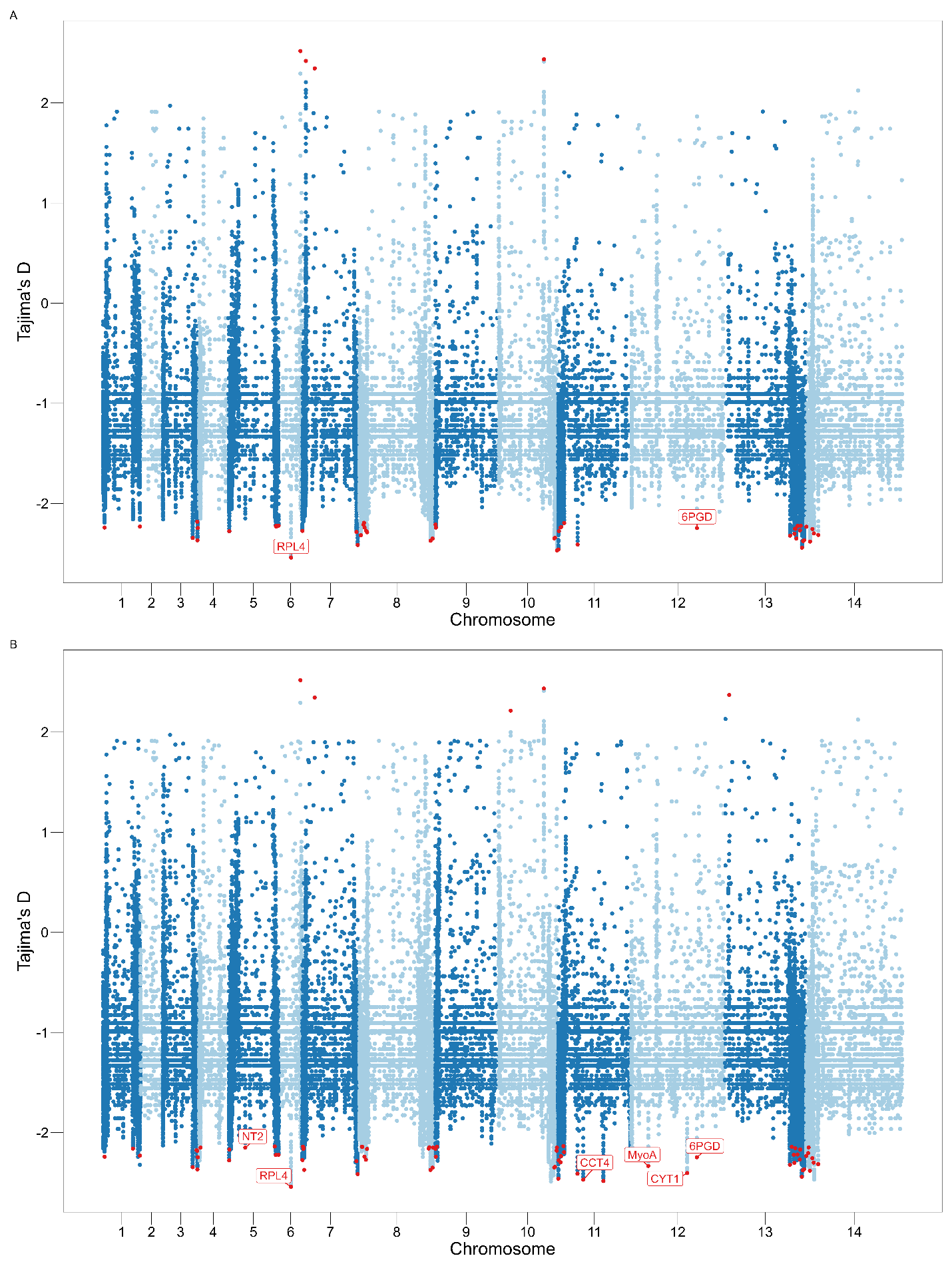

**Supplemental Figure 8 – Tajima’s D values in 300 bp windows across A) genes and B) exons.** The top values of |D| are highlighted in red and those with known gene annotations are labeled. Only PmmyoA has a known function relevant to RBC invasion. No putative antimalarial resistance genes are highlighted.

**Supplemental Table 1 – Likelihood and fit of demographic models**

| **Model** | **Log Likelihood** | **CL-AIC** |
| --- | --- | --- |
| Bottlegrowth | -556 | 14,058 |
| Growth | -868 | -1,204 |
| Standard Neutral Model | -46,556 | NA |
| Two Epoch | -911 | 27,976 |
| Three Epoch | -514 | -59,633 |

**Supplemental Table 2 – Genome-wide Tajima’s D top hits.** “Where Found” column reflects whether the gene in question was a top hit for gene scans, exon scans, or both.

| **CHROM** | **Gene ID** | **Gene Name** | **Description** | **Biotype** | **Where Found** | **Largest Tajima Value** | **Absolute Tajima** | **Selection Type** |
| --- | --- | --- | --- | --- | --- | --- | --- | --- |
| PmUG01_06_v1 | PmUG01_06017800 | RPL4 | 60S ribosomal protein L4%2C putative | protein_coding | Both | -2.54 | 2.54 | Directional |
| PmUG01_06_v1 | PmUG01_06025500 |  | Plasmodium exported protein%2C unknown function | protein_coding | Both | 2.52 | 2.52 | Balancing |
| PmUG01_11_v1 | PmUG01_11044200 |  | cell division cycle protein 48 homologue%2C putative | protein_coding | Exons | -2.48 | 2.48 | Directional |
| PmUG01_10_v1 | PmUG01_10054800 |  | fam-l protein | protein_coding | Both | -2.47 | 2.47 | Directional |
| PmUG01_11_v1 | PmUG01_11028200 | CCT4 | T-complex protein 1%2C delta subunit%2C putative | protein_coding | Exons | -2.47 | 2.47 | Directional |
| PmUG01_11_v1 | PmUG01_11010600 |  | Plasmodium exported protein%2C unknown function | protein_coding | Both | -2.46 | 2.46 | Directional |
| PmUG01_13_v1 | PmUG01_13065600 |  | Plasmodium exported protein%2C unknown function | protein_coding | Both | -2.44 | 2.44 | Directional |
| PmUG01_10_v1 | PmUG01_10046600 |  | hypothetical protein | protein_coding | Both | 2.44 | 2.44 | Balancing |
| PmUG01_07_v1 | PmUG01_07012200 |  | STP1 protein | protein_coding | Genes | 2.42 | 2.42 | Balancing |
| PmUG01_07_v1 | PmUG01_07051600 |  | Plasmodium exported protein%2C unknown function | protein_coding | Both | -2.41 | 2.41 | Directional |
| PmUG01_11_v1 | PmUG01_11024100 |  | cytochrome c oxidase subunit ApiCOX24%2C putative | protein_coding | Both | -2.41 | 2.41 | Directional |
| PmUG01_12_v1 | PmUG01_12054200 | CYT1 | cytochrome c1%2C heme protein%2C mitochondrial%2C putative | protein_coding | Exons | -2.40 | 2.40 | Directional |
| PmUG01_14_v1 | PmUG01_14012400 |  | fam-m protein | protein_coding | Both | -2.38 | 2.38 | Directional |
| PmUG01_13_v1 | PmUG01_13066500 |  | Plasmodium exported protein%2C unknown function | protein_coding | Both | -2.38 | 2.38 | Directional |
| PmUG01_08_v1 | PmUG01_08061200 |  | STP1 protein | protein_coding | Both | -2.37 | 2.37 | Directional |
| PmUG01_07_v1 | PmUG01_07011200 |  | fam-l protein | protein_coding | Exons | -2.37 | 2.37 | Directional |
| PmUG01_13_v1 | PmUG01_13012600 |  | dynein heavy chain%2C putative | protein_coding | Exons | 2.37 | 2.37 | Balancing |
| PmUG01_03_v1 | PmUG01_03035500 |  | fam-m protein | protein_coding | Both | -2.37 | 2.37 | Directional |
| PmUG01_13_v1 | PmUG01_13067400 |  | fam-l protein | protein_coding | Both | -2.37 | 2.37 | Directional |
| PmUG01_08_v1 | PmUG01_08062600 |  | Plasmodium exported protein%2C unknown function | protein_coding | Both | -2.35 | 2.35 | Directional |
| PmUG01_13_v1 | PmUG01_13061600 |  | fam-m protein | protein_coding | Genes | -2.35 | 2.35 | Directional |
| PmUG01_10_v1 | PmUG01_10053100 |  | fam-m protein | protein_coding | Both | -2.35 | 2.35 | Directional |
| PmUG01_07_v1 | PmUG01_07017300 |  | TFIIS domain-containing protein | protein_coding | Both | 2.35 | 2.35 | Balancing |
| PmUG01_03_v1 | PmUG01_03032200 |  | hypothetical protein | protein_coding | Both | -2.34 | 2.34 | Directional |
| PmUG01_10_v1 | PmUG01_10053500 |  | fam-l protein | protein_coding | Both | -2.34 | 2.34 | Directional |
| PmUG01_12_v1 | PmUG01_12022300 | MyoA | myosin A%2C putative | protein_coding | Exons | -2.33 | 2.33 | Directional |
| PmUG01_13_v1 | PmUG01_13057500 |  | Plasmodium exported protein%2C unknown function | protein_coding | Both | -2.32 | 2.32 | Directional |
| PmUG01_08_v1 | PmUG01_08011800 |  | fam-l protein | protein_coding | Genes | -2.32 | 2.32 | Directional |
| PmUG01_14_v1 | PmUG01_14017600 |  | gamete antigen 27/25%2C putative | protein_coding | Both | -2.31 | 2.31 | Directional |
| PmUG01_13_v1 | PmUG01_13060300 |  | fam-m protein | protein_coding | Both | -2.30 | 2.30 | Directional |
| PmUG01_14_v1 | PmUG01_14014500 |  | Plasmodium exported protein%2C unknown function | protein_coding | Both | -2.30 | 2.30 | Directional |
| PmUG01_08_v1 | PmUG01_08016300 |  | STP1 protein | protein_coding | Both | -2.29 | 2.29 | Directional |
| PmUG01_07_v1 | PmUG01_07050700 |  | fam-l protein | protein_coding | Both | -2.28 | 2.28 | Directional |
| PmUG01_05_v1 | PmUG01_05011100 |  | Plasmodium exported protein%2C unknown function | protein_coding | Both | -2.28 | 2.28 | Directional |
| PmUG01_05_v1 | PmUG01_05011000 |  | Plasmodium exported protein%2C unknown function | protein_coding | Both | -2.28 | 2.28 | Directional |
| PmUG01_11_v1 | PmUG01_11010900 |  | STP1 protein | protein_coding | Both | -2.28 | 2.28 | Directional |
| PmUG01_07_v1 | PmUG01_07010200 |  | fam-l protein | protein_coding | Both | -2.27 | 2.27 | Directional |
| PmUG01_08_v1 | PmUG01_08015400 |  | fam-l protein | protein_coding | Both | -2.27 | 2.27 | Directional |
| PmUG01_13_v1 | PmUG01_13064400 |  | Plasmodium exported protein%2C unknown function | protein_coding | Both | -2.27 | 2.27 | Directional |
| PmUG01_14_v1 | PmUG01_14013800 |  | Plasmodium exported protein%2C unknown function | protein_coding | Both | -2.26 | 2.26 | Directional |
| PmUG01_13_v1 | PmUG01_13061000 |  | fam-l protein | protein_coding | Both | -2.25 | 2.25 | Directional |
| PmUG01_13_v1 | PmUG01_13060600 |  | fam-m protein | protein_coding | Genes | -2.25 | 2.25 | Directional |
| PmUG01_12_v1 | PmUG01_12062300 | 6PGD | 6-phosphogluconate dehydrogenase%2C decarboxylating%2C putative | protein_coding | Both | -2.25 | 2.25 | Directional |
| PmUG01_03_v1 | PmUG01_03035700 |  | Plasmodium exported protein%2C unknown function | protein_coding | Both | -2.25 | 2.25 | Directional |
| PmUG01_09_v1 | PmUG01_09010500 |  | fam-m protein | protein_coding | Both | -2.24 | 2.24 | Directional |
| PmUG01_01_v1 | PmUG01_01011400 |  | fam-m protein | protein_coding | Both | -2.24 | 2.24 | Directional |
| PmUG01_08_v1 | PmUG01_08014300 |  | fam-m protein | protein_coding | Both | -2.24 | 2.24 | Directional |
| PmUG01_11_v1 | PmUG01_11012000 |  | fam-m protein | protein_coding | Both | -2.24 | 2.24 | Directional |
| PmUG01_11_v1 | PmUG01_11012200 |  | fam-l protein | protein_coding | Both | -2.24 | 2.24 | Directional |
| PmUG01_14_v1 | PmUG01_14010200 |  | Plasmodium exported protein%2C unknown function | protein_coding | Both | -2.23 | 2.23 | Directional |
| PmUG01_05_v1 | PmUG01_05043400 |  | fam-l protein | protein_coding | Genes | -2.23 | 2.23 | Directional |
| PmUG01_01_v1 | PmUG01_01034800 |  | fam-l protein | protein_coding | Both | -2.23 | 2.23 | Directional |
| PmUG01_13_v1 | PmUG01_13062700 |  | fam-m protein | protein_coding | Both | -2.22 | 2.22 | Directional |
| PmUG01_05_v1 | PmUG01_05043300 |  | fam-l protein | protein_coding | Both | -2.22 | 2.22 | Directional |
| PmUG01_13_v1 | PmUG01_13064800 |  | fam-m protein | protein_coding | Genes | -2.22 | 2.22 | Directional |
| PmUG01_05_v1 | PmUG01_05045000 |  | fam-m protein | protein_coding | Both | -2.22 | 2.22 | Directional |
| PmUG01_13_v1 | PmUG01_13060000 |  | fam-m protein | protein_coding | Exons | -2.22 | 2.22 | Directional |
| PmUG01_10_v1 | PmUG01_10020700 |  | MerC domain-containing protein%2C putative | protein_coding | Exons | 2.21 | 2.21 | Balancing |
| PmUG01_08_v1 | PmUG01_08013700 |  | fam-m protein | protein_coding | Genes | -2.21 | 2.21 | Directional |
| PmUG01_09_v1 | PmUG01_09010300 |  | Plasmodium exported protein%2C unknown function | protein_coding | Both | -2.21 | 2.21 | Directional |
| PmUG01_14_v1 | PmUG01_14011200 |  | fam-m protein | protein_coding | Exons | -2.20 | 2.20 | Directional |
| PmUG01_11_v1 | PmUG01_11013900 |  | fam-l protein | protein_coding | Both | -2.20 | 2.20 | Directional |
| PmUG01_08_v1 | PmUG01_08014000 |  | fam-l protein | protein_coding | Genes | -2.19 | 2.19 | Directional |
| PmUG01_03_v1 | PmUG01_03035400 |  | fam-l protein | protein_coding | Both | -2.18 | 2.18 | Directional |
| PmUG01_10_v1 | PmUG01_10054600 |  | Plasmodium exported protein%2C unknown function | protein_coding | Exons | -2.18 | 2.18 | Directional |
| PmUG01_03_v1 | PmUG01_03035100 |  | fam-l protein | protein_coding | Exons | -2.18 | 2.18 | Directional |
| PmUG01_13_v1 | PmUG01_13064600 |  | fam-l protein | protein_coding | Exons | -2.17 | 2.17 | Directional |
| PmUG01_07_v1 | PmUG01_07010800 |  | fam-l protein | protein_coding | Exons | -2.16 | 2.16 | Directional |
| PmUG01_01_v1 | PmUG01_01031000 |  | Plasmodium exported protein%2C unknown function | protein_coding | Exons | -2.16 | 2.16 | Directional |
| PmUG01_08_v1 | PmUG01_08060200 |  | Plasmodium exported protein%2C unknown function | protein_coding | Exons | -2.16 | 2.16 | Directional |
| PmUG01_08_v1 | PmUG01_08062900 |  | fam-l protein | protein_coding | Exons | -2.15 | 2.15 | Directional |
| PmUG01_05_v1 | PmUG01_05021600 | NT2 | nucleoside transporter 2%2C putative | protein_coding | Exons | -2.15 | 2.15 | Directional |
| PmUG01_04_v1 | PmUG01_04011200 |  | fam-m protein | protein_coding | Exons | -2.15 | 2.15 | Directional |
| PmUG01_14_v1 | PmUG01_14011600 |  | fam-l protein | protein_coding | Exons | -2.15 | 2.15 | Directional |
| PmUG01_07_v1 | PmUG01_07010700 |  | fam-m protein | protein_coding | Exons | -2.15 | 2.15 | Directional |
| PmUG01_08_v1 | PmUG01_08060600 |  | fam-m protein | protein_coding | Exons | -2.15 | 2.15 | Directional |
| PmUG01_08_v1 | PmUG01_08013100 |  | Plasmodium exported protein%2C unknown function | protein_coding | Exons | -2.14 | 2.14 | Directional |
| PmUG01_13_v1 | PmUG01_13058600 |  | Plasmodium exported protein%2C unknown function | protein_coding | Exons | -2.14 | 2.14 | Directional |
| PmUG01_08_v1 | PmUG01_08012600 |  | fam-l protein | protein_coding | Exons | -2.14 | 2.14 | Directional |
| PmUG01_07_v1 | PmUG01_07010500 |  | fam-m protein | protein_coding | Exons | -2.14 | 2.14 | Directional |
| PmUG01_09_v1 | PmUG01_09011200 |  | Plasmodium exported protein%2C unknown function | protein_coding | Exons | -2.14 | 2.14 | Directional |
| PmUG01_05_v1 | PmUG01_05042500 |  | Plasmodium exported protein%2C unknown function | protein_coding | Exons | -2.14 | 2.14 | Directional |
| PmUG01_11_v1 | PmUG01_11013500 |  | Plasmodium exported protein%2C unknown function | protein_coding | Exons | -2.13 | 2.13 | Directional |

**Supplemental Table 3 – Genome-wide nS_L_ top hits with a minor allele frequency cutoff of 0.05 applied**

| **CHROM** | **Gene ID** | **Gene Name** | **Description** | **Biotype** | **Largest *n*S_L_ Value** | **Absolute *n*S_L_** |
| --- | --- | --- | --- | --- | --- | --- |
| PmUG01_01_v1 | PmUG01_01011800 |  | fam-l protein | protein_coding | -2.95 | 2.95 |
| PmUG01_01_v1 | PmUG01_01012600 |  | STP1 protein | protein_coding | 3.00 | 3.00 |
| PmUG01_01_v1 | PmUG01_01028700 |  | filament assembling protein%2C putative | protein_coding | 3.13 | 3.13 |
| PmUG01_01_v1 | PmUG01_01028800 | UTP14 | U3 small nucleolar RNA-associated protein 14%2C putative | protein_coding | 2.86 | 2.86 |
| PmUG01_01_v1 | PmUG01_01029300 | ACbeta | adenylyl cyclase beta%2C putative | protein_coding | 3.10 | 3.10 |
| PmUG01_01_v1 | PmUG01_01034500 |  | Plasmodium exported protein%2C unknown function | protein_coding | -3.40 | 3.40 |
| PmUG01_05_v1 | PmUG01_05011800 |  | fam-l protein | protein_coding | 2.80 | 2.80 |
| PmUG01_05_v1 | PmUG01_05016400 | ETRAMP | early transcribed membrane protein | protein_coding | 2.70 | 2.70 |
| PmUG01_05_v1 | PmUG01_05026000 |  | RNA-binding protein%2C putative | protein_coding | 2.81 | 2.81 |
| PmUG01_05_v1 | PmUG01_05026600 |  | conserved protein%2C unknown function | protein_coding | 2.72 | 2.72 |
| PmUG01_05_v1 | PmUG01_05027600 |  | Plasmodium exported protein (PHIST)%2C unknown function | protein_coding | 3.83 | 3.83 |
| PmUG01_05_v1 | PmUG01_05029000 |  | GTP-binding protein%2C putative | protein_coding | 3.51 | 3.51 |
| PmUG01_05_v1 | PmUG01_05032900 | CZIF2 | C3H1-type zinc finger protein CZIF2%2C putative | protein_coding | 3.05 | 3.05 |
| PmUG01_05_v1 | PmUG01_05034400 |  | conserved Plasmodium protein%2C unknown function | protein_coding | 2.69 | 2.69 |
| PmUG01_05_v1 | PmUG01_05034900 |  | conserved Plasmodium protein%2C unknown function | protein_coding | 2.81 | 2.81 |
| PmUG01_05_v1 | PmUG01_05035500 |  | conserved Plasmodium protein%2C unknown function | protein_coding | 2.82 | 2.82 |
| PmUG01_05_v1 | PmUG01_05039800 |  | BSD-domain protein%2C putative | protein_coding | 2.88 | 2.88 |
| PmUG01_08_v1 | PmUG01_08033400 |  | conserved Plasmodium protein%2C unknown function | protein_coding | 2.79 | 2.79 |
| PmUG01_08_v1 | PmUG01_08038400 |  | CPSF (cleavage and polyadenylation specific factor)%2C subunit A%2C putative | protein_coding | 2.72 | 2.72 |
| PmUG01_08_v1 | PmUG01_08039200 |  | E3 ubiquitin-protein ligase%2C putative | protein_coding | 3.06 | 3.06 |
| PmUG01_08_v1 | PmUG01_08042600 |  | conserved Plasmodium protein%2C unknown function | protein_coding | 3.24 | 3.24 |
| PmUG01_08_v1 | PmUG01_08048500 | AKIT10 | apicomplexan kinetochore protein 10%2C putative | protein_coding | 2.78 | 2.78 |
| PmUG01_08_v1 | PmUG01_08050100 |  | conserved Plasmodium protein%2C unknown function | protein_coding | 2.84 | 2.84 |
| PmUG01_09_v1 | PmUG01_09010500 |  | fam-m protein | protein_coding | -2.91 | 2.91 |
| PmUG01_09_v1 | PmUG01_09010600 |  | Plasmodium exported protein%2C unknown function | protein_coding | -2.91 | 2.91 |
| PmUG01_10_v1 | PmUG01_10037200 |  | conserved protein%2C unknown function | protein_coding | 2.83 | 2.83 |
| PmUG01_10_v1 | PmUG01_10038200 |  | conserved Plasmodium protein%2C unknown function | protein_coding | 3.04 | 3.04 |
| PmUG01_10_v1 | PmUG01_10046700 |  | merozoite surface protein%2C putative | protein_coding | 2.73 | 2.73 |
| PmUG01_10_v1 | PmUG01_10053800 |  | Plasmodium exported protein%2C unknown function | protein_coding | -3.04 | 3.04 |
| PmUG01_11_v1 | PmUG01_11012100 |  | fam-l protein | protein_coding | -2.91 | 2.91 |
| PmUG01_14_v1 | PmUG01_14076000 |  | rab specific GDP dissociation inhibitor%2C putative | protein_coding | 3.02 | 3.02 |

**Supplemental Table 4 – Genome-wide nS_L_ top hits with no minor allele frequency cutoff applied**

| **CHROM** | **Gene ID** | **Gene Name** | **Description** | **Biotype** | **Largest *n*S_L_ Value** | **Absolute *n*S_L_** |
| --- | --- | --- | --- | --- | --- | --- |
| PmUG01_01_v1 | PmUG01_01010200 |  | fam-m protein | protein_coding | -4.51 | 4.51 |
| PmUG01_01_v1 | PmUG01_01011800 |  | fam-l protein | protein_coding | -5.14 | 5.14 |
| PmUG01_01_v1 | PmUG01_01012600 |  | STP1 protein | protein_coding | 3.52 | 3.52 |
| PmUG01_01_v1 | PmUG01_01021000 | MED14 | mediator of RNA polymerase II transcription subunit 14%2C putative | protein_coding | 3.17 | 3.17 |
| PmUG01_01_v1 | PmUG01_01021800 |  | Sfi1-like protein SLP%2C putative | protein_coding | -4.09 | 4.09 |
| PmUG01_01_v1 | PmUG01_01026700 |  | zinc finger protein%2C putative | protein_coding | -3.65 | 3.65 |
| PmUG01_01_v1 | PmUG01_01028700 |  | filament assembling protein%2C putative | protein_coding | -4.16 | 4.16 |
| PmUG01_01_v1 | PmUG01_01033700 |  | fam-l protein | protein_coding | 3.75 | 3.75 |
| PmUG01_01_v1 | PmUG01_01034500 |  | Plasmodium exported protein%2C unknown function | protein_coding | -4.18 | 4.18 |
| PmUG01_02_v1 | PmUG01_02015100 |  | mitochondrial carrier protein%2C putative | protein_coding | -3.21 | 3.21 |
| PmUG01_02_v1 | PmUG01_02015200 | PSOP24 | secreted ookinete protein%2C putative | protein_coding | -3.50 | 3.50 |
| PmUG01_02_v1 | PmUG01_02020500 | VPS51 | vacuolar protein sorting-associated protein 51%2C putative | protein_coding | 5.19 | 5.19 |
| PmUG01_02_v1 | PmUG01_02020700 |  | aspartyl-tRNA synthetase%2C putative | protein_coding | 3.62 | 3.62 |
| PmUG01_02_v1 | PmUG01_02021000 |  | conserved Plasmodium membrane protein%2C unknown function | protein_coding | -3.35 | 3.35 |
| PmUG01_03_v1 | PmUG01_03031000 |  | protein kinase%2C putative | protein_coding | 3.25 | 3.25 |
| PmUG01_03_v1 | PmUG01_03033700 |  | fam-l protein | protein_coding | -3.26 | 3.26 |
| PmUG01_03_v1 | PmUG01_03035400 |  | fam-l protein | protein_coding | -3.14 | 3.14 |
| PmUG01_03_v1 | PmUG01_03035700 |  | Plasmodium exported protein%2C unknown function | protein_coding | -3.45 | 3.45 |
| PmUG01_03_v1 | PmUG01_03035900 |  | fam-m protein | protein_coding | -3.30 | 3.30 |
| PmUG01_04_v1 | PmUG01_04011300 |  | STP1 protein | protein_coding | -3.25 | 3.25 |
| PmUG01_04_v1 | PmUG01_04017400 | RON6 | rhoptry neck protein 6%2C putative | protein_coding | -3.30 | 3.30 |
| PmUG01_04_v1 | PmUG01_04018700 |  | conserved Plasmodium protein%2C unknown function | protein_coding | -4.12 | 4.12 |
| PmUG01_04_v1 | PmUG01_04024600 |  | serine-repeat antigen%2C putative | protein_coding | -3.12 | 3.12 |
| PmUG01_04_v1 | PmUG01_04026400 | RAD2 | DNA repair protein RAD2%2C putative | protein_coding | -3.14 | 3.14 |
| PmUG01_05_v1 | PmUG01_05010700 |  | fam-l protein | protein_coding | -4.02 | 4.02 |
| PmUG01_05_v1 | PmUG01_05011100 |  | Plasmodium exported protein%2C unknown function | protein_coding | -3.80 | 3.80 |
| PmUG01_05_v1 | PmUG01_05011800 |  | fam-l protein | protein_coding | 4.12 | 4.12 |
| PmUG01_05_v1 | PmUG01_05016500 |  | conserved Plasmodium protein%2C unknown function | protein_coding | -3.68 | 3.68 |
| PmUG01_05_v1 | PmUG01_05018100 | PDI8 | protein disulfide-isomerase%2C putative | protein_coding | -3.82 | 3.82 |
| PmUG01_05_v1 | PmUG01_05018800 |  | conserved Plasmodium protein%2C unknown function | protein_coding | -3.80 | 3.80 |
| PmUG01_05_v1 | PmUG01_05019400 |  | SNARE protein%2C putative | protein_coding | -3.26 | 3.26 |
| PmUG01_05_v1 | PmUG01_05021600 | NT2 | nucleoside transporter 2%2C putative | protein_coding | -3.53 | 3.53 |
| PmUG01_05_v1 | PmUG01_05027600 |  | Plasmodium exported protein (PHIST)%2C unknown function | protein_coding | -3.73 | 3.73 |
| PmUG01_05_v1 | PmUG01_05027700 |  | RNA-binding protein%2C putative | protein_coding | -3.69 | 3.69 |
| PmUG01_05_v1 | PmUG01_05030200 | PGM2 | phosphoglucomutase-2%2C putative | protein_coding | -3.17 | 3.17 |
| PmUG01_05_v1 | PmUG01_05030600 | Ub | ubiquitin%2C putative | protein_coding | -3.76 | 3.76 |
| PmUG01_05_v1 | PmUG01_05035200 |  | conserved Plasmodium protein%2C unknown function | protein_coding | -3.35 | 3.35 |
| PmUG01_05_v1 | PmUG01_05036800 |  | conserved Plasmodium protein%2C unknown function | protein_coding | -3.97 | 3.97 |
| PmUG01_05_v1 | PmUG01_05043300 |  | fam-l protein | protein_coding | -4.43 | 4.43 |
| PmUG01_05_v1 | PmUG01_05043700 |  | fam-l protein | protein_coding | -3.65 | 3.65 |
| PmUG01_05_v1 | PmUG01_05044200 |  | fam-m protein | protein_coding | -3.90 | 3.90 |
| PmUG01_05_v1 | PmUG01_05044900 |  | fam-l protein | protein_coding | -4.20 | 4.20 |
| PmUG01_05_v1 | PmUG01_05045000 |  | fam-m protein | protein_coding | -4.22 | 4.22 |
| PmUG01_06_v1 | PmUG01_06013000 | DPA | deoxyribose-phosphate aldolase%2C putative | protein_coding | -4.01 | 4.01 |
| PmUG01_06_v1 | PmUG01_06016800 | SET9 | SET domain protein%2C putative | protein_coding | 3.28 | 3.28 |
| PmUG01_06_v1 | PmUG01_06020600 |  | conserved Plasmodium protein%2C unknown function | protein_coding | -3.89 | 3.89 |
| PmUG01_06_v1 | PmUG01_06021600 |  | merozoite surface protein 3%2C putative | protein_coding | -3.48 | 3.48 |
| PmUG01_06_v1 | PmUG01_06021900 |  | merozoite surface protein 3%2C putative | protein_coding | 3.88 | 3.88 |
| PmUG01_06_v1 | PmUG01_06022900 |  | merozoite surface protein 3%2C putative | protein_coding | 4.00 | 4.00 |
| PmUG01_06_v1 | PmUG01_06024300 | VPS11 | vacuolar protein sorting-associated protein 11%2C putative | protein_coding | -3.84 | 3.84 |
| PmUG01_06_v1 | PmUG01_06025600 |  | Plasmodium exported protein%2C unknown function | protein_coding | -3.50 | 3.50 |
| PmUG01_07_v1 | PmUG01_07010800 |  | fam-l protein | protein_coding | -3.54 | 3.54 |
| PmUG01_07_v1 | PmUG01_07011200 |  | fam-l protein | protein_coding | -3.55 | 3.55 |
| PmUG01_07_v1 | PmUG01_07011300 |  | fam-l protein | protein_coding | -3.44 | 3.44 |
| PmUG01_07_v1 | PmUG01_07012200 |  | STP1 protein | protein_coding | -3.13 | 3.13 |
| PmUG01_07_v1 | PmUG01_07050800 |  | fam-m protein | protein_coding | -3.60 | 3.60 |
| PmUG01_07_v1 | PmUG01_07051400 |  | fam-l protein | protein_coding | -3.60 | 3.60 |
| PmUG01_07_v1 | PmUG01_07051600 |  | Plasmodium exported protein%2C unknown function | protein_coding | -3.56 | 3.56 |
| PmUG01_08_v1 | PmUG01_08011700 |  | fam-m protein | protein_coding | -3.57 | 3.57 |
| PmUG01_08_v1 | PmUG01_08011800 |  | fam-l protein | protein_coding | -3.75 | 3.75 |
| PmUG01_08_v1 | PmUG01_08012800 |  | fam-l protein | protein_coding | -3.54 | 3.54 |
| PmUG01_08_v1 | PmUG01_08013700 |  | fam-m protein | protein_coding | -3.58 | 3.58 |
| PmUG01_08_v1 | PmUG01_08014000 |  | fam-l protein | protein_coding | -3.53 | 3.53 |
| PmUG01_08_v1 | PmUG01_08014300 |  | fam-m protein | protein_coding | -4.08 | 4.08 |
| PmUG01_08_v1 | PmUG01_08015400 |  | fam-l protein | protein_coding | -3.67 | 3.67 |
| PmUG01_08_v1 | PmUG01_08044900 |  | phosphatidylinositol 3- and 4-kinase%2C putative | protein_coding | 3.14 | 3.14 |
| PmUG01_08_v1 | PmUG01_08046900 |  | N2227-like protein%2C putative | protein_coding | 3.31 | 3.31 |
| PmUG01_08_v1 | PmUG01_08056400 |  | Plasmodium exported protein%2C unknown function | protein_coding | -3.14 | 3.14 |
| PmUG01_08_v1 | PmUG01_08057000 |  | fam-m protein | protein_coding | -3.67 | 3.67 |
| PmUG01_08_v1 | PmUG01_08057700 |  | fam-l protein | protein_coding | -3.62 | 3.62 |
| PmUG01_08_v1 | PmUG01_08060200 |  | Plasmodium exported protein%2C unknown function | protein_coding | -3.16 | 3.16 |
| PmUG01_08_v1 | PmUG01_08060500 |  | fam-l protein | protein_coding | -3.25 | 3.25 |
| PmUG01_08_v1 | PmUG01_08060800 |  | fam-l protein | protein_coding | -3.61 | 3.61 |
| PmUG01_08_v1 | PmUG01_08062600 |  | Plasmodium exported protein%2C unknown function | protein_coding | -3.27 | 3.27 |
| PmUG01_08_v1 | PmUG01_08062900 |  | fam-l protein | protein_coding | -3.53 | 3.53 |
| PmUG01_08_v1 | PmUG01_08063800 |  | fam-l protein | protein_coding | -3.12 | 3.12 |
| PmUG01_09_v1 | PmUG01_09010300 |  | Plasmodium exported protein%2C unknown function | protein_coding | -3.77 | 3.77 |
| PmUG01_09_v1 | PmUG01_09010600 |  | Plasmodium exported protein%2C unknown function | protein_coding | -3.91 | 3.91 |
| PmUG01_09_v1 | PmUG01_09025200 | DHHC9 | palmitoyltransferase DHHC9%2C putative | protein_coding | -4.16 | 4.16 |
| PmUG01_09_v1 | PmUG01_09031700 |  | protein KIC10%2C putative | protein_coding | -3.41 | 3.41 |
| PmUG01_09_v1 | PmUG01_09052600 |  | regulator of chromosome condensation%2C putative | protein_coding | -4.25 | 4.25 |
| PmUG01_09_v1 | PmUG01_09055000 | RSA4 | ribosome assembly protein 4%2C putative | protein_coding | -3.96 | 3.96 |
| PmUG01_10_v1 | PmUG01_10013600 | FRM1 | formin 1%2C putative | protein_coding | 3.46 | 3.46 |
| PmUG01_10_v1 | PmUG01_10019600 | TYW1 | S-adenosyl-L-methionine-dependent tRNA 4-demethylwyosine synthase%2C putative | protein_coding | -3.14 | 3.14 |
| PmUG01_10_v1 | PmUG01_10040200 |  | conserved Plasmodium protein%2C unknown function | protein_coding | -3.15 | 3.15 |
| PmUG01_10_v1 | PmUG01_10046700 |  | merozoite surface protein%2C putative | protein_coding | 3.25 | 3.25 |
| PmUG01_10_v1 | PmUG01_10053100 |  | fam-m protein | protein_coding | -4.22 | 4.22 |
| PmUG01_10_v1 | PmUG01_10053800 |  | Plasmodium exported protein%2C unknown function | protein_coding | -3.45 | 3.45 |
| PmUG01_10_v1 | PmUG01_10054000 |  | Plasmodium exported protein%2C unknown function | protein_coding | -3.97 | 3.97 |
| PmUG01_10_v1 | PmUG01_10054500 |  | Plasmodium exported protein%2C unknown function | protein_coding | -3.21 | 3.21 |
| PmUG01_10_v1 | PmUG01_10054800 |  | fam-l protein | protein_coding | -3.62 | 3.62 |
| PmUG01_10_v1 | PmUG01_10054900 |  | fam-m protein | protein_coding | -3.23 | 3.23 |
| PmUG01_11_v1 | PmUG01_11012100 |  | fam-l protein | protein_coding | -3.36 | 3.36 |
| PmUG01_11_v1 | PmUG01_11013600 |  | fam-l protein | protein_coding | -3.60 | 3.60 |
| PmUG01_11_v1 | PmUG01_11053000 |  | tetratricopeptide repeat protein%2C putative | protein_coding | -3.21 | 3.21 |
| PmUG01_11_v1 | PmUG01_11056900 |  | elongation of fatty acids protein%2C putative | protein_coding | 3.40 | 3.40 |
| PmUG01_12_v1 | PmUG01_12010600 |  | Plasmodium exported protein%2C unknown function | protein_coding | -3.25 | 3.25 |
| PmUG01_12_v1 | PmUG01_12016600 |  | elongation factor Tu%2C putative | protein_coding | -3.32 | 3.32 |
| PmUG01_12_v1 | PmUG01_12019900 |  | RING zinc finger protein%2C putative | protein_coding | -3.94 | 3.94 |
| PmUG01_12_v1 | PmUG01_12020600 |  | zinc finger protein%2C putative | protein_coding | -3.91 | 3.91 |
| PmUG01_12_v1 | PmUG01_12030200 |  | MSP7-like protein%2C putative | protein_coding | 3.37 | 3.37 |
| PmUG01_12_v1 | PmUG01_12032300 | TRAPPC2 | trafficking protein particle complex subunit 2%2C putative | protein_coding | 3.24 | 3.24 |
| PmUG01_12_v1 | PmUG01_12033500 |  | helicase%2C putative | protein_coding | -3.99 | 3.99 |
| PmUG01_12_v1 | PmUG01_12044800 | YIP1 | protein transport protein YIP1%2C putative | protein_coding | -3.81 | 3.81 |
| PmUG01_12_v1 | PmUG01_12048900 |  | WD repeat-containing protein%2C putative | protein_coding | -4.09 | 4.09 |
| PmUG01_12_v1 | PmUG01_12054300 |  | conserved Plasmodium protein%2C unknown function | protein_coding | -3.89 | 3.89 |
| PmUG01_12_v1 | PmUG01_12055100 |  | conserved Plasmodium protein%2C unknown function | protein_coding | -3.55 | 3.55 |
| PmUG01_12_v1 | PmUG01_12058600 |  | alpha/beta hydrolase%2C putative | protein_coding | -4.06 | 4.06 |
| PmUG01_12_v1 | PmUG01_12062500 | ISCU | iron-sulfur cluster assembly protein ISCU%2C putative | protein_coding | 3.16 | 3.16 |
| PmUG01_12_v1 | PmUG01_12062600 | APP | aminopeptidase P%2C putative | protein_coding | 3.36 | 3.36 |
| PmUG01_12_v1 | PmUG01_12069200 |  | calponin homology domain-containing protein%2C putative | protein_coding | -3.91 | 3.91 |
| PmUG01_12_v1 | PmUG01_12079700 |  | proteasome subunit alpha type-1%2C putative | protein_coding | -3.71 | 3.71 |
| PmUG01_13_v1 | PmUG01_13016100 | ISN1 | IMP-specific 5'-nucleotidase 1%2C putative | protein_coding | -3.17 | 3.17 |
| PmUG01_13_v1 | PmUG01_13051300 | RlmN | radical SAM protein%2C putative | protein_coding | -3.28 | 3.28 |
| PmUG01_13_v1 | PmUG01_13053100 | BET5 | trafficking protein particle complex subunit 1%2C putative | protein_coding | -3.22 | 3.22 |
| PmUG01_13_v1 | PmUG01_13054500 | MISFIT | nuclear formin-like protein MISFIT%2C putative | protein_coding | -3.29 | 3.29 |
| PmUG01_13_v1 | PmUG01_13054900 |  | dynein regulatory complex protein%2C putative | protein_coding | -3.34 | 3.34 |
| PmUG01_13_v1 | PmUG01_13055800 |  | ribosomal protein S27a%2C putative | protein_coding | -3.23 | 3.23 |
| PmUG01_13_v1 | PmUG01_13057500 |  | Plasmodium exported protein%2C unknown function | protein_coding | -3.59 | 3.59 |
| PmUG01_13_v1 | PmUG01_13058800 |  | Plasmodium exported protein%2C unknown function | protein_coding | -3.66 | 3.66 |
| PmUG01_13_v1 | PmUG01_13059600 |  | fam-m protein | protein_coding | -3.45 | 3.45 |
| PmUG01_13_v1 | PmUG01_13062700 |  | fam-m protein | protein_coding | -3.72 | 3.72 |
| PmUG01_13_v1 | PmUG01_13064400 |  | Plasmodium exported protein%2C unknown function | protein_coding | -3.62 | 3.62 |
| PmUG01_13_v1 | PmUG01_13064600 |  | fam-l protein | protein_coding | -3.30 | 3.30 |
| PmUG01_13_v1 | PmUG01_13064700 |  | fam-l protein | protein_coding | -3.41 | 3.41 |
| PmUG01_13_v1 | PmUG01_13064800 |  | fam-m protein | protein_coding | -3.63 | 3.63 |
| PmUG01_13_v1 | PmUG01_13068500 |  | fam-m protein | protein_coding | -3.39 | 3.39 |
| PmUG01_14_v1 | PmUG01_14011600 |  | fam-l protein | protein_coding | -3.41 | 3.41 |
| PmUG01_14_v1 | PmUG01_14012600 |  | fam-m protein | protein_coding | -3.42 | 3.42 |
| PmUG01_14_v1 | PmUG01_14013100 |  | fam-m protein | protein_coding | -3.59 | 3.59 |
| PmUG01_14_v1 | PmUG01_14013400 |  | fam-m protein | protein_coding | -3.28 | 3.28 |
| PmUG01_14_v1 | PmUG01_14024900 |  | carbamoyl phosphate synthetase%2C putative | protein_coding | -3.37 | 3.37 |
| PmUG01_14_v1 | PmUG01_14036100 |  | conserved protein%2C unknown function | protein_coding | -3.14 | 3.14 |
| PmUG01_14_v1 | PmUG01_14036300 |  | female development protein FD3%2C putative | protein_coding | -3.31 | 3.31 |
| PmUG01_14_v1 | PmUG01_14051900 | TOP6A | meiotic recombination protein SPO11%2C putative | protein_coding | -3.37 | 3.37 |
| PmUG01_14_v1 | PmUG01_14076000 |  | rab specific GDP dissociation inhibitor%2C putative | protein_coding | 3.45 | 3.45 |
| PmUG01_14_v1 | PmUG01_14078500 | UBC12 | NEDD8-conjugating enzyme UBC12%2C putative | protein_coding | -3.41 | 3.41 |
